# Clinical outcomes of Lassa fever in West Africa: A systematic review and meta-analysis

**DOI:** 10.1101/2025.01.10.24319564

**Authors:** Azuka Patrick Okwuraiwe, Chizaram Anselm Onyeaghala, Obiageli Theresa Ozoude, Muritala Odidi Suleiman, Samirah Nndwan Abdu-Aguye, Nkolika Jacinta Ezekwelu, Tolulope Amos Oyeniyi, Ayodapo Oluwadare Jegede, Adaeze Elfrida Egwudo, Oluchukwu Perpetua Okeke, Olunuke Rebecca Abodunrin, Folahanmi Tomiwa Akinsolu, Olajide Odunayo Sobande

## Abstract

**Introduction:** Lassa fever is an acute viral haemorrhagic fever that poses a substantial public health security threat in West Africa. Due to its non-specific clinical manifestations and the absence of a reliable point-of-care test, diagnosis could be delayed, leading to severe complications and mortality during epidemics. This systematic review aimed to determine the clinical outcomes of LF in West Africa.

**Methods:** A systematic review and meta-analyses were performed by conducting an extensive online search using PubMed, Web of Science, Scopus, CINAHL, and Google Scholar (PROSPERO protocol CRD42024587426). Only articles written in English were included in publications from 2014 to 2024. The analysis followed PRISMA guidelines. The mortality rate of LF was pooled using a random effects model.

**Results:** We included 19 studies that contained data from 4177 patients hospitalized with LF of any age. Most of the studies were of retrospective cohort (16/19; 84.2%) study design and were predominantly conducted in Nigeria (16/19; 84.2%). The mortality rate was highest in a Sierra Leonean study (63.0%), whereas group-based analysis among the Nigerian studies identified Owo as having the highest mortality rate of 13% (95% CI: 06-23; I^2^ =98%). The pooled LF mortality rate was 19% (95% confidence interval [CI]:10-32). The most common complications of LF are acute kidney injury (AKI) at a pooled proportion of 19% (95% CI; 13-26; I^2^=89%)), followed by abnormal bleeding at a pooled proportion of 17% (95% CI; 9-30; I^2^=98%), and CNS manifestations at a pooled proportion of 15% (95% CI; 6-32; I^2^=98%)).

**Conclusion:** With one out of every five hospitalized Lassa Fever patients likely to die in West Africa, accelerating the development of rapid diagnostic tests, licensed vaccines, and novel therapeutics is crucial. Strengthening community engagement, risk communication, and healthcare worker training will enhance early diagnosis and effective case management thereby reducing mortality and severe complications.

## Introduction

Lassa fever (LF) is an acute viral haemorrhagic fever (VHF) that poses a significant threat to health security. First described in 1969 in Nigeria, Lassa fever’s causative agent is the Lassa virus (LASV), a member of the Arenaviridae family and a biosafety level 4 pathogen. It is endemic in parts of West Africa, including Nigeria, Sierra Leone, Guinea, and Liberia.[1] The main reservoir and primary source of infection is the multimammate mouse (*Mastomys natalensis*). [2] However, the virus also spreads between human beings via contact with the body fluids of an infected person, consumption, or handling of contaminated food or household items. An estimated 300,000-500,000 people are infected with LASV every year in West Africa, although the overall incidence is likely to be underestimated with 5,000-10,000 deaths annually. [3]

Nigeria has the highest reported cases of LF estimated at 25% of the total disease burden. [4] The incubation period of Lassa fever is 3-21 days. [5] Nigeria has experienced increases in confirmed cases and deaths yearly since 2017, although this may be attributed to heightened clinical awareness and improvements in polymerase chain reaction-based diagnostic capacity. Lassa fever is said to have seasonal transmission annually between November and March, however, there are increasing reports of double peaks due to climate change (rainfall and increased temperature) related influence on the multiplicity of the rodent vector and the emergence of new hosts (*Mastomys erythroleucus* and *Hylomyscus pamfi*). [6,7] Due to the epidemic potential of Lassa fever and its high case fatality rates (CFR) in hospitalized patients, Lassa fever was included in the priority list of diseases for research and development, urging intensified research, including improved treatments, a blueprint of the World Health Organization. [8–10] The only known treatment is the broad-spectrum antiviral drug ribavirin, shown to be most effective in the first six days, after the onset of symptoms. No approved vaccine is currently available for LASV, although progress has been made in the multiagency Coalition for Epidemic Preparedness Innovations-funded Lassa fever (IAVI-C105) clinical trials [11,12] and with human monoclonal antibodies and other antivirals in preclinical animal models. [13,14]

Most individuals infected with LASV in Africa are never diagnosed with Lassa fever because of mild or asymptomatic presentation of the disease and the lack of readily available diagnostic assays. [15] Clinical manifestations are variable, and often non-specific as many of those are seen in other febrile infectious diseases in endemic areas such as malaria, and typhoid fever. Although the overall case fatality rate (CFR) is 1%, between 10% and 20% of patients treated in hospitals with Lassa fever die, which can be much higher (above 40%) in outbreak settings or individuals at increased risk. Outbreaks frequently occur in Nigeria and 38% of cases were fatal in the outbreak of 2015. [15] Available studies show a wide spectrum of clinical severities, ranging from asymptomatic infection to serious multiorgan dysfunction and death. Even among acute cases, observations range from minimal cell damage in the liver and kidneys to more extensive involvement of these organs. [15] Severe Lassa fever with a lethal outcome is associated with encephalopathy, acute kidney injury, respiratory failure, and death.

This systematic review, therefore, sets out to determine the clinical outcomes of Lassa fever in West Africa. This is aimed at providing useful information that will improve early diagnosis and clinical management of Lassa fever in West Africa.

## Methods

### Protocol registration

The study protocol was prospectively registered in the International Prospective Register of Systematic Review, PROSPERO (CRD42024587426). The conduct and reporting of this systematic review and meta-analysis were guided by the Preferred Reporting Items for Systematic Reviews and Meta-Analyses (PRISMA). [16]

### Search strategy

An extensive online search was conducted using four top academic databases for biomedical journals; PubMed, Web of Science, Scopus, CINAHL, and Google Scholar. The search strategy included peer-reviewed studies published between 1 September 2004 and 31 August 2024. In addition, the reference lists of included studies were screened to identify additional publications.

The search string included combinations of the following 3 keywords: terms related to “clinical outcomes”, “Lassa fever”, “West Africa”. Appropriate medical subject headings (MeSH) terms were added to the databases and this was adjusted to optimize the search for relevant literature. The Boolean operators or connector words “AND” and “OR” were used to combine or exclude search terms effectively and achieve a final result set. All identified articles were imported into Rayyan software for de-duplication and to organize the bibliography. In addition to keeping track of search resources, dates resources searched were noted to ensure that the review included the most current evidence-based literature on the review question.

## Review Questions

### Primary Outcomes

1. What is the number of participants with mortality for Lassa fever infection.
2. What is the number of participants who recovered from Lassa fever infection
3. What is the number of participants who developed complications due to Lassa fever infection.
4. What is the number of participants who were lost to follow-up.

### Inclusion and exclusion Criteria

All studies that reported clinical outcomes of Lassa fever in West Africa, in the English language, and published between January 2014 and December 2024 were eligible. Quantitative studies such as cohort studies, retrospective studies, case-control studies, and interventional studies that discussed the recovery, mortality, complications, and loss to follow-up of Lassa fever patients were included in the review. Studies were screened using the PICOTS framework (Population, Intervention, Comparators, Outcomes, Time, Studies).

Studies were excluded if they were found to contain only abstracts or were cross-sectional studies, case reports, and case series.

### Data extraction

Data extraction was done by two independent reviewers (O.T.O., and M.O.S., with discrepancies resolved by a third reviewer A.P.O.) using a pretested data extraction form prepared in Microsoft Excel. Reliability between reviewers was ensured by using the Kappa scores. The reviewers independently extracted relevant information, including first author, publication year, country (city/area) of study, sample size, gender, study design, inclusion criteria, number of participants confirmed for Lassa fever infection, clinical outcomes such as recovery, mortality, mean age, age range, complications and lost to follow-up.

### Selection of studies

We downloaded all titles and abstracts retrieved by electronic searching to the reference management software Zotero version 6.0.23. We removed duplicates, and three review authors (O.T.O., M.O.S., and A.P.O) independently examined the remaining references. We excluded studies that did not meet the inclusion criteria and obtained copies of the full text of potentially relevant studies. Two review authors (O.T.O and M.O.S.) independently assessed the eligibility of the retrieved papers and resolved any disagreements by discussion or recourse to a third review author (A.P.O. or C.A.O.).

### Methodological quality and risk of bias assessment

The methodology quality of the included studies was assessed using the Critical Appraisal tools for use in the Joanna Briggs Institute (JBI) Systematic Reviews Checklist for cohort studies. [17] Two independent reviewers assessed the methodological quality and risk of bias using the JBI checklist for cohort studies to assess for risk of bias. Discrepancies were resolved by a third reviewer. Eleven quality domains were used for this assessment. They included the description of similarity among the two groups recruited, the similarity of measured exposures, validity, and reliability of measured exposures, identification of confounding factors, strategies to deal with confounding factors, being free of the outcome at the start or the moment of exposure, valid and reliable measurement of outcomes, sufficient time for report and manifestations of outcomes, completeness or non-completeness of follow up, strategies to addressing incomplete follow up, and use of appropriate statistical analysis tool. In addition, a unique identification was assigned to each selected study. The total score ranged from 0 to 11, with the overall score categorized as follows: 0–3: “high risk,” 4–7: “moderate risk,” and 8–11: “low risk” of bias.

### Statistical analysis

A meta-analysis was performed on studies reporting clinical outcomes of Lassa fever, including death, survival, abnormal bleeding, acute kidney disease, and central nervous system (CNS) manifestations. The pooled proportions for each clinical outcome were calculated along with 95% confidence intervals (CIs). Statistical heterogeneity among studies was assessed using Cochran’s Q test and the I² statistic. A P-value of <0.1 for the Q test and an I² value greater than 50% indicated significant heterogeneity. Given the expected variability, a random-effects model was employed to pool outcomes, yielding a more conservative estimate of prevalence.

Additionally, subgroup analyses were conducted based on the publication year of the included studies. For studies conducted in Nigeria, further subgroup analyses were performed based on the specific locations of these studies.

Funnel plots were utilized to evaluate publication bias for the primary outcome. A leave-one-out analysis was executed to systematically exclude each study, assessing the robustness of the results and the influence of individual studies on the pooled prevalence.

All statistical analyses were conducted using R software (version 4.4.2).

## Results

### Selection of studies

A total of 2022 records were retrieved through electronic searches, of which 476 were from electronically published databases, and 1546 from other search engines (Google Scholar). After removing 409 duplicate records, 1613 remained from which 1275 were excluded following title and abstract screening. The full-text records for the remaining 338 studies were obtained for detailed evaluations, after which 281 studies were excluded during resolution of conflict. In addition, only studies that showed clear methodology were included. Finally, 19 met the study inclusion criteria. The full search summary is presented in the Transparent Reporting of Systematic Reviews and Meta-analyses (PRISMA) Flow Diagram in Figure 1.

**Figure 1:**
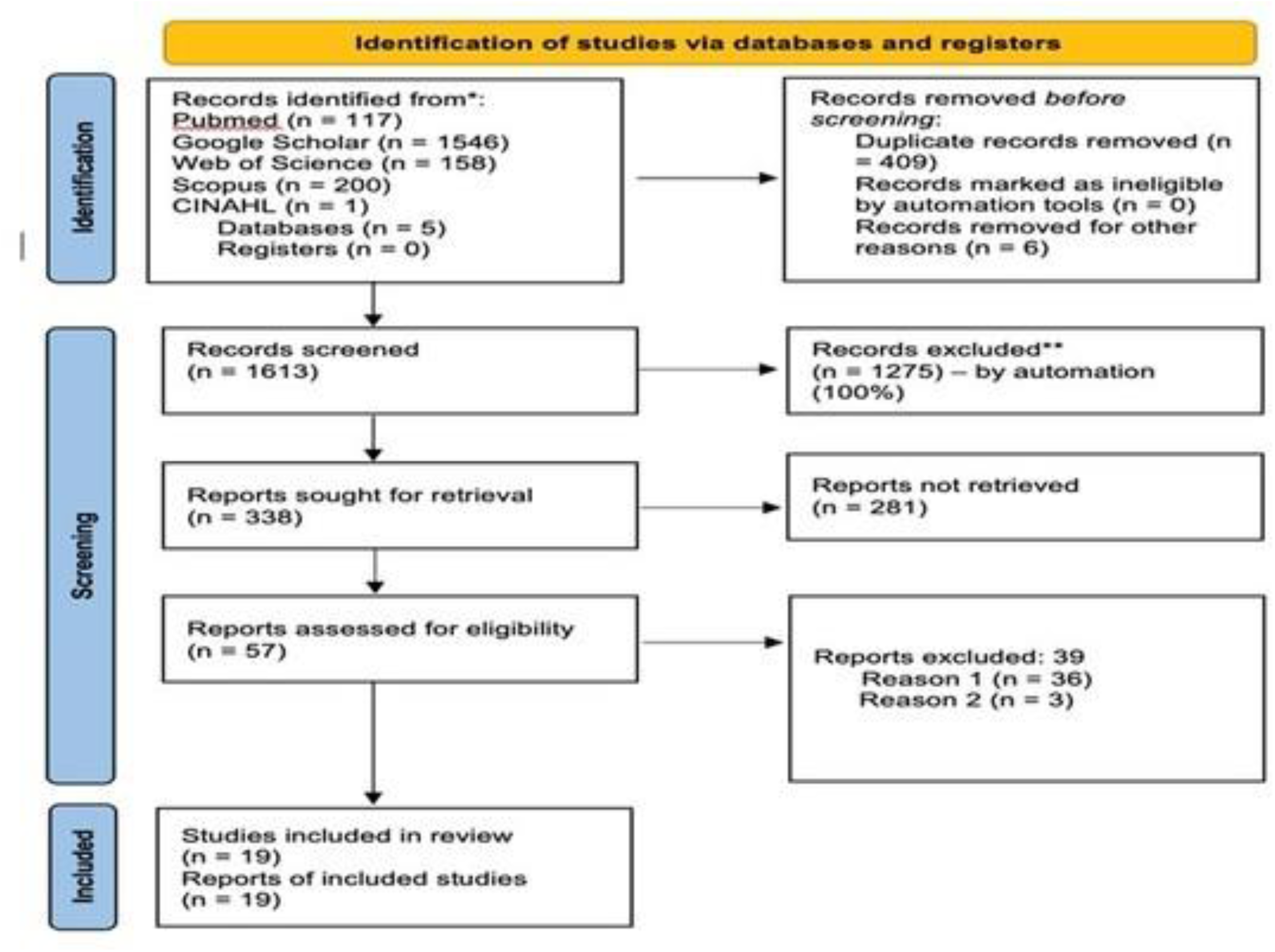
PRISMA flow chart.

### Characteristics of included studies

A total of 4177 subjects were covered by the 19 synthesized studies, with the sample size in individual studies ranging from 11 to 1594, a median of 100, and a mean of 219.8.

The age range of the enrolled subjects was provided by 84.2% of the studies with the widest age range being 1–90 years while the narrowest was 5–12 years. Some studies did not report the mean or median and age range of the sample. However, 4/19 studies (12.8%) did not report any mean or median age data. Many of the studies reported gender-aggregated data.

Synthesized studies were predominantly Nigerian studies [see Figure 2], (16/19; 84.2%), in Owo (6/16; 37.5%), and of retrospective cohort (16/19; 84.2%) study design. The earliest study was conducted in 2014 and the latest in 2024 (Table 1). The number of articles published was highest in 2020 (Figure 3).

**Figure 2:**
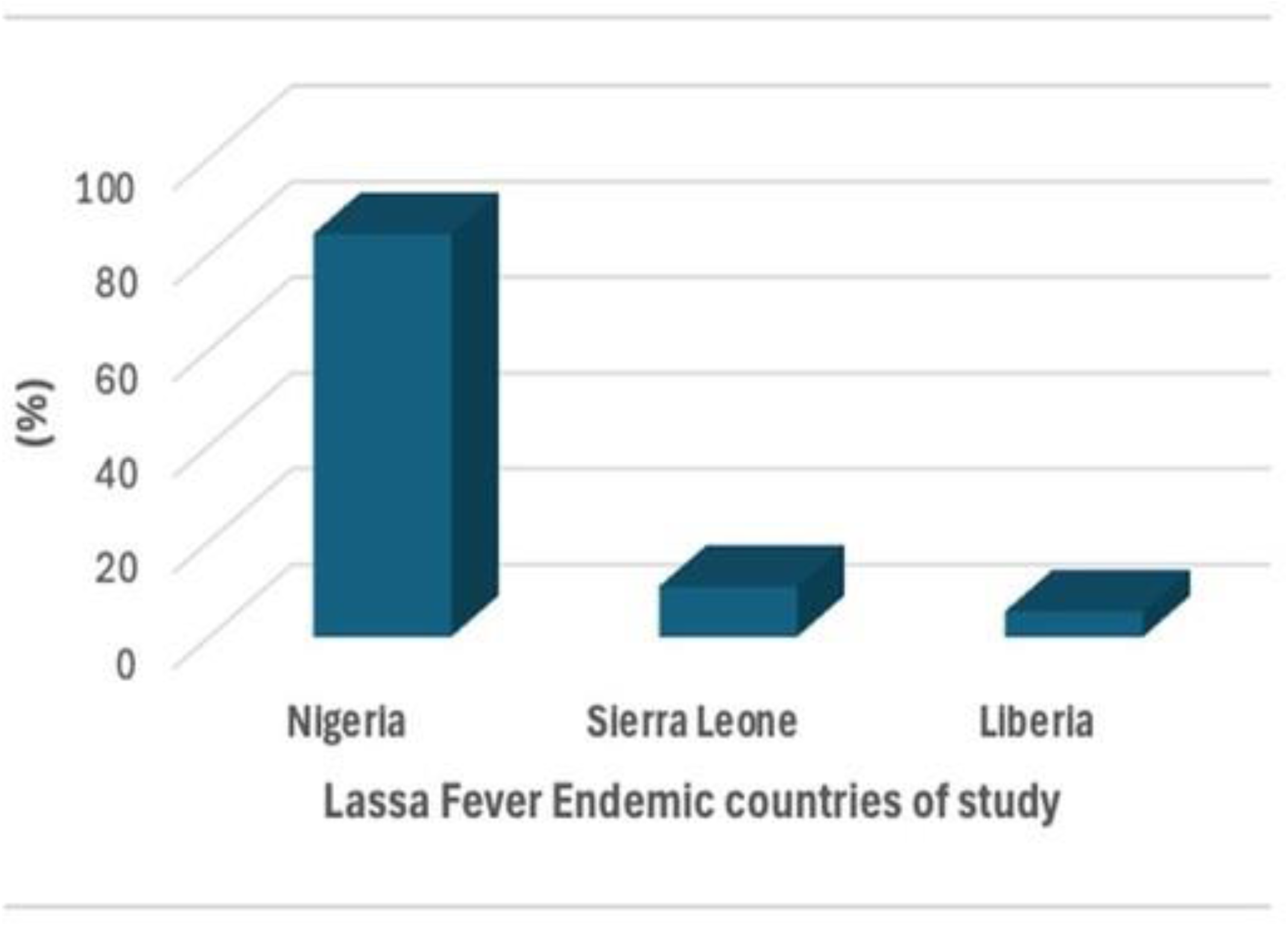
Countries that reported clinical outcomes of Lassa fever in West Africa.

**Figure 3:**
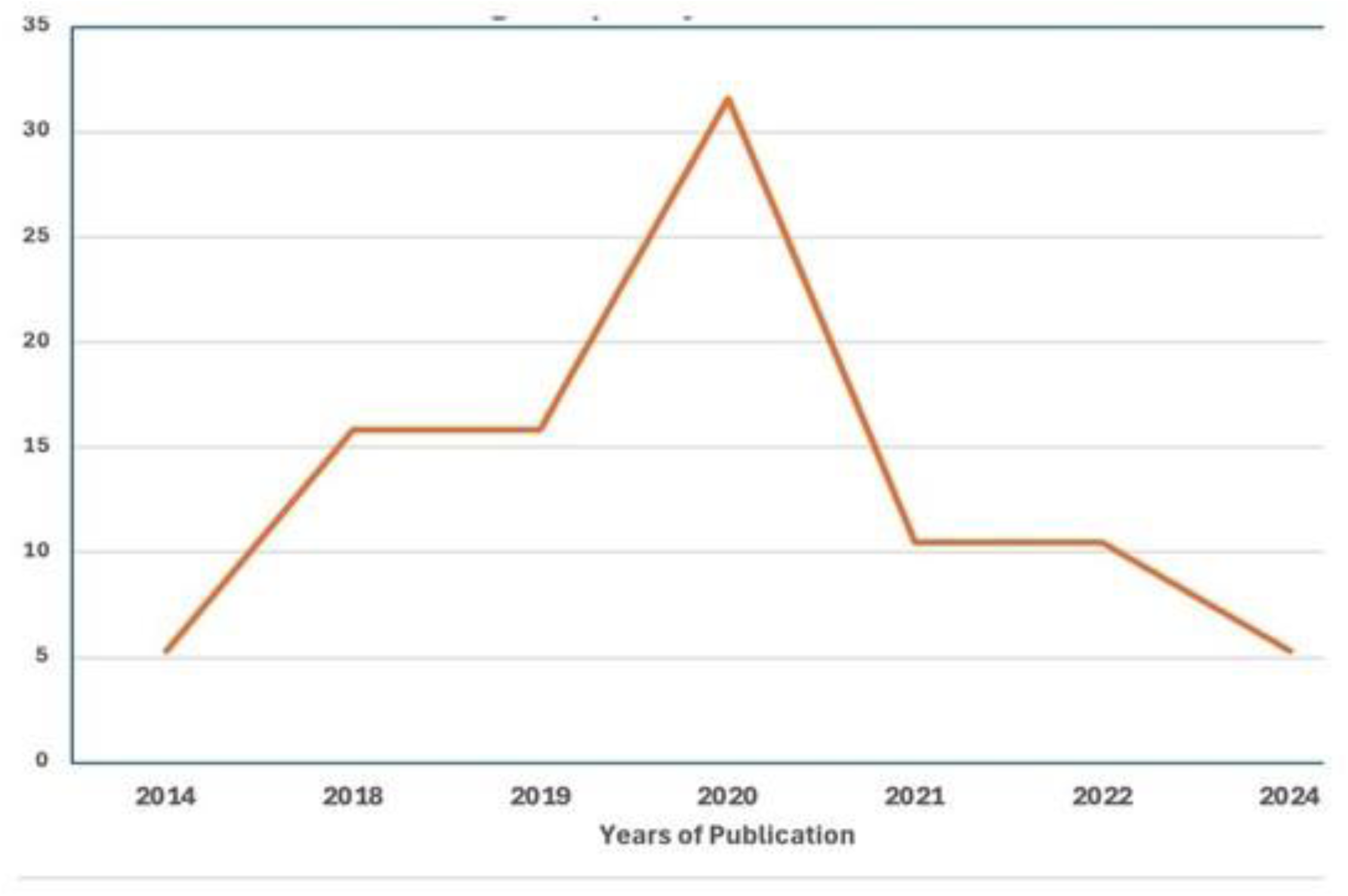
Frequency of included studies based on year of publication.

**Table 1:**
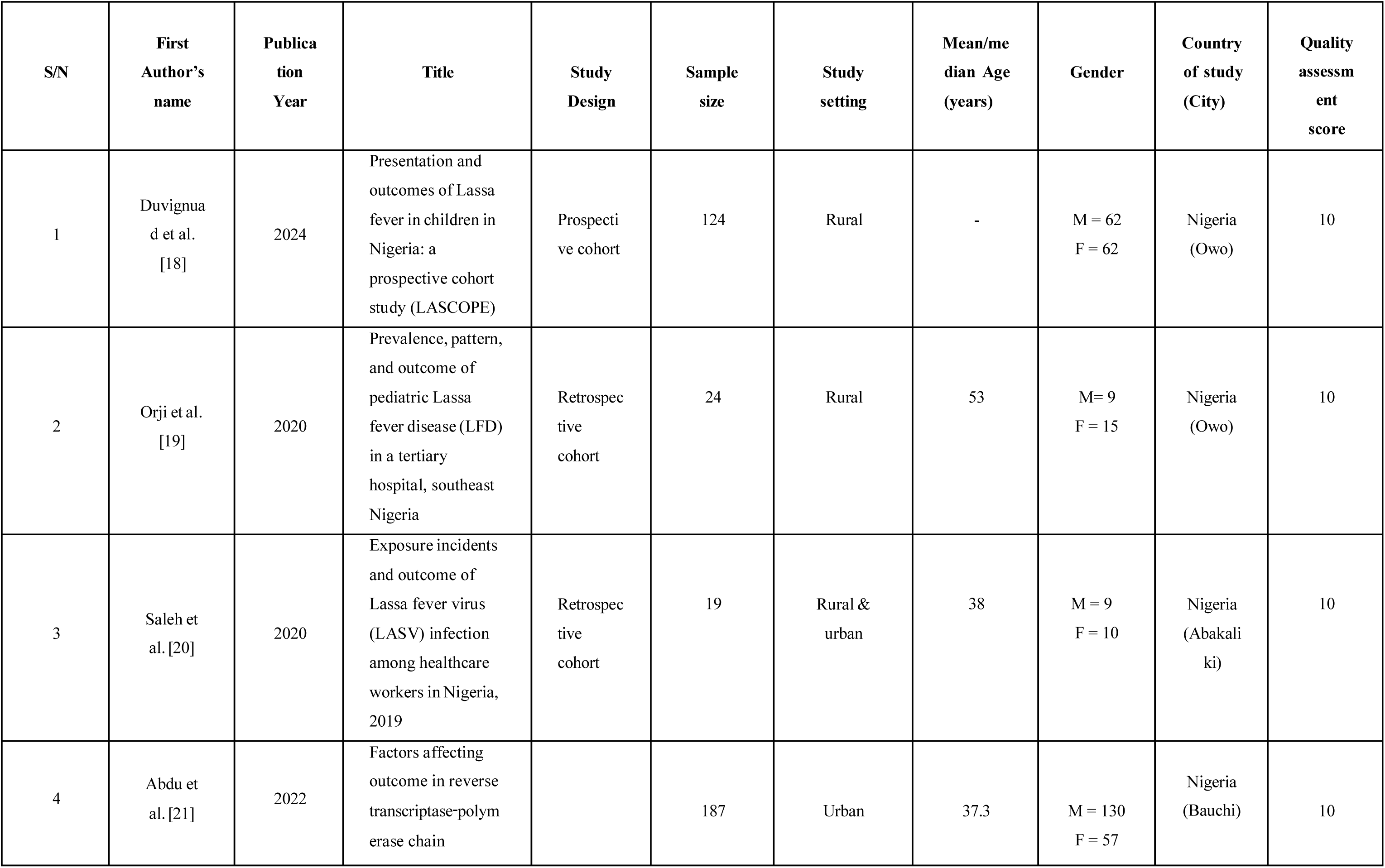
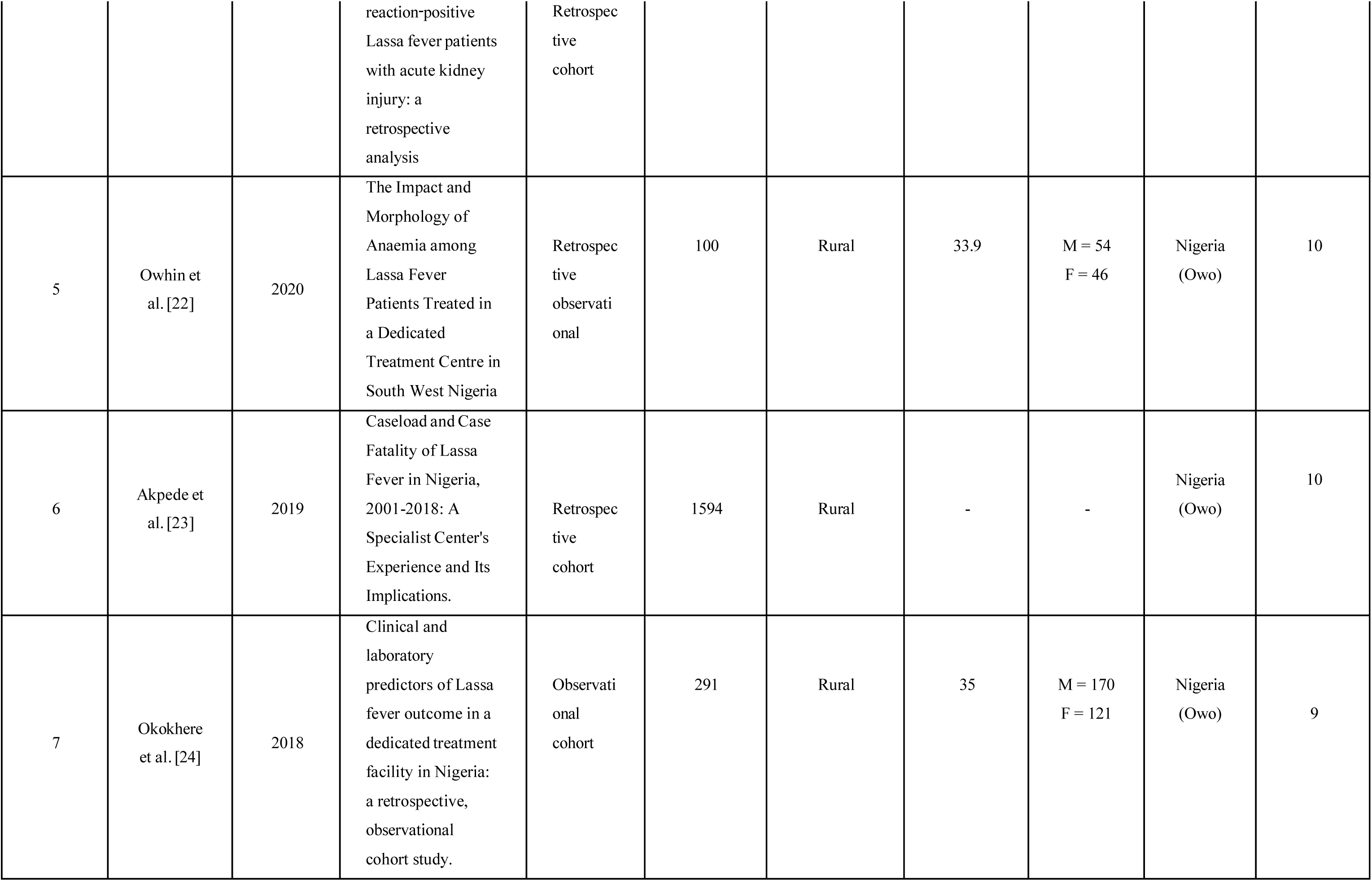
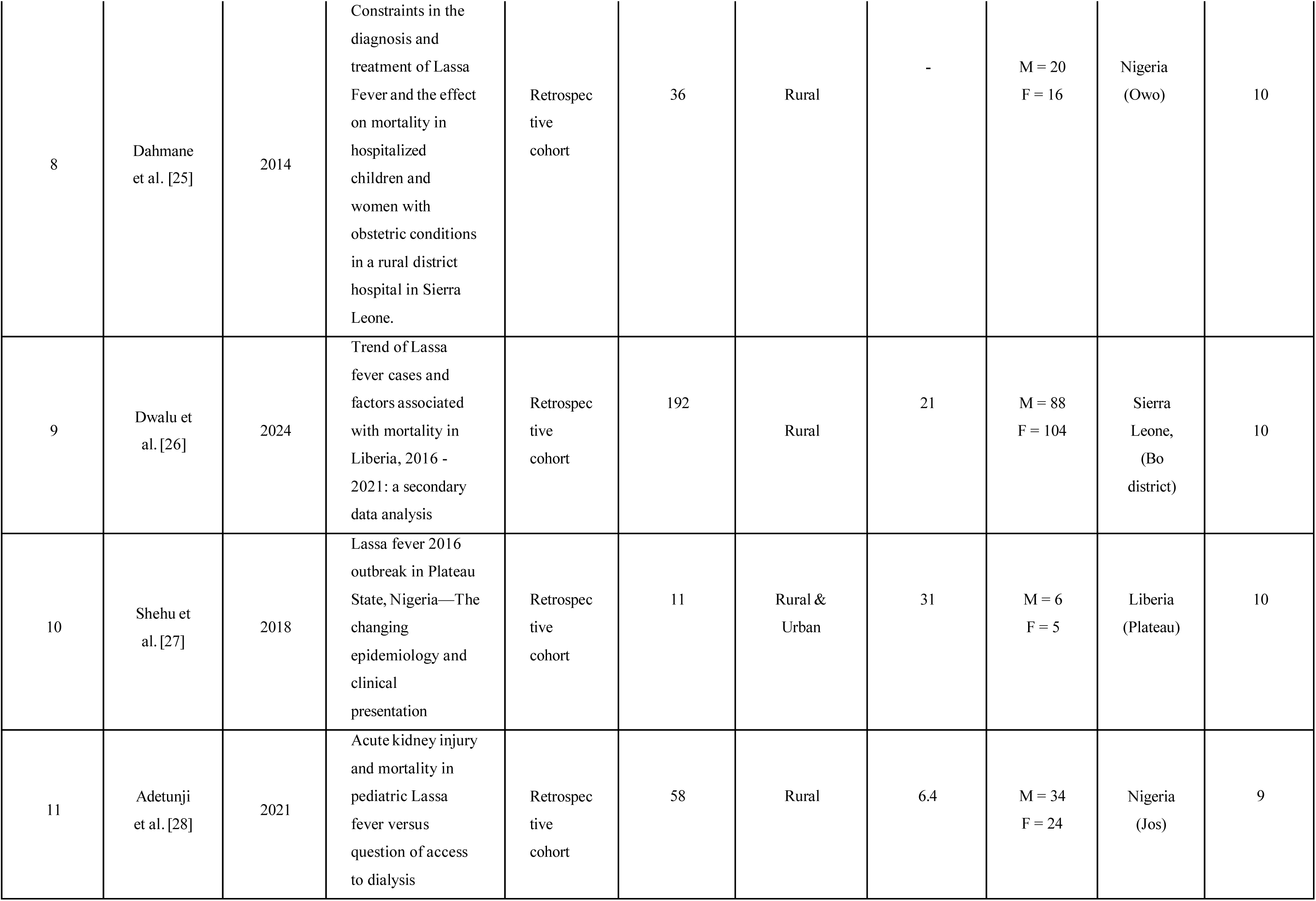
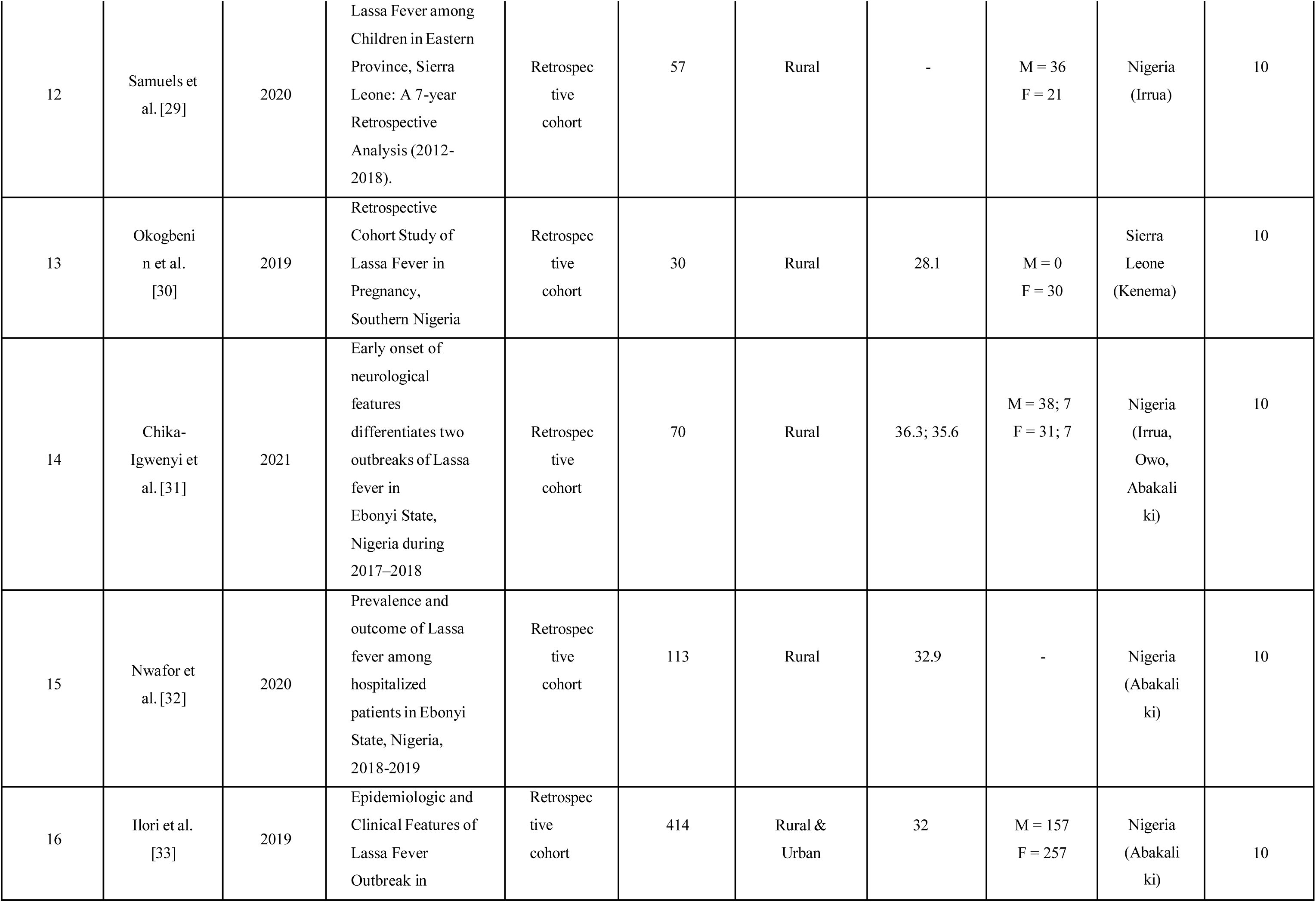
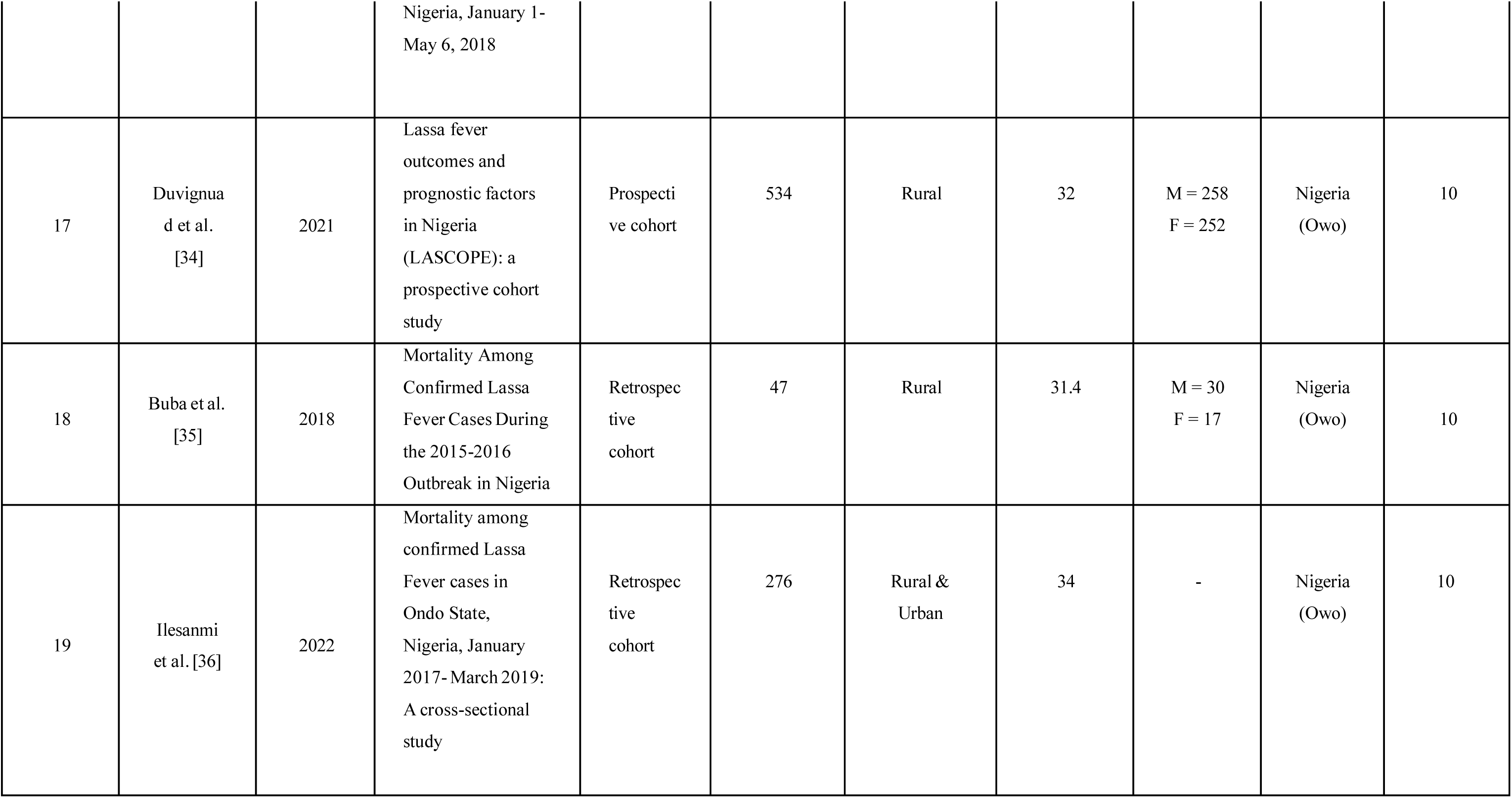
Characteristics of included studies.

### Primary outcomes

Recovery: Among synthesized studies, recovery rates ranged from 37% to 96.8% (Table 2). The recovery rate was highest in the prospective cohort study conducted by Duvignaud and colleagues in 2024 (96.8%). [34]

**Table 2:**
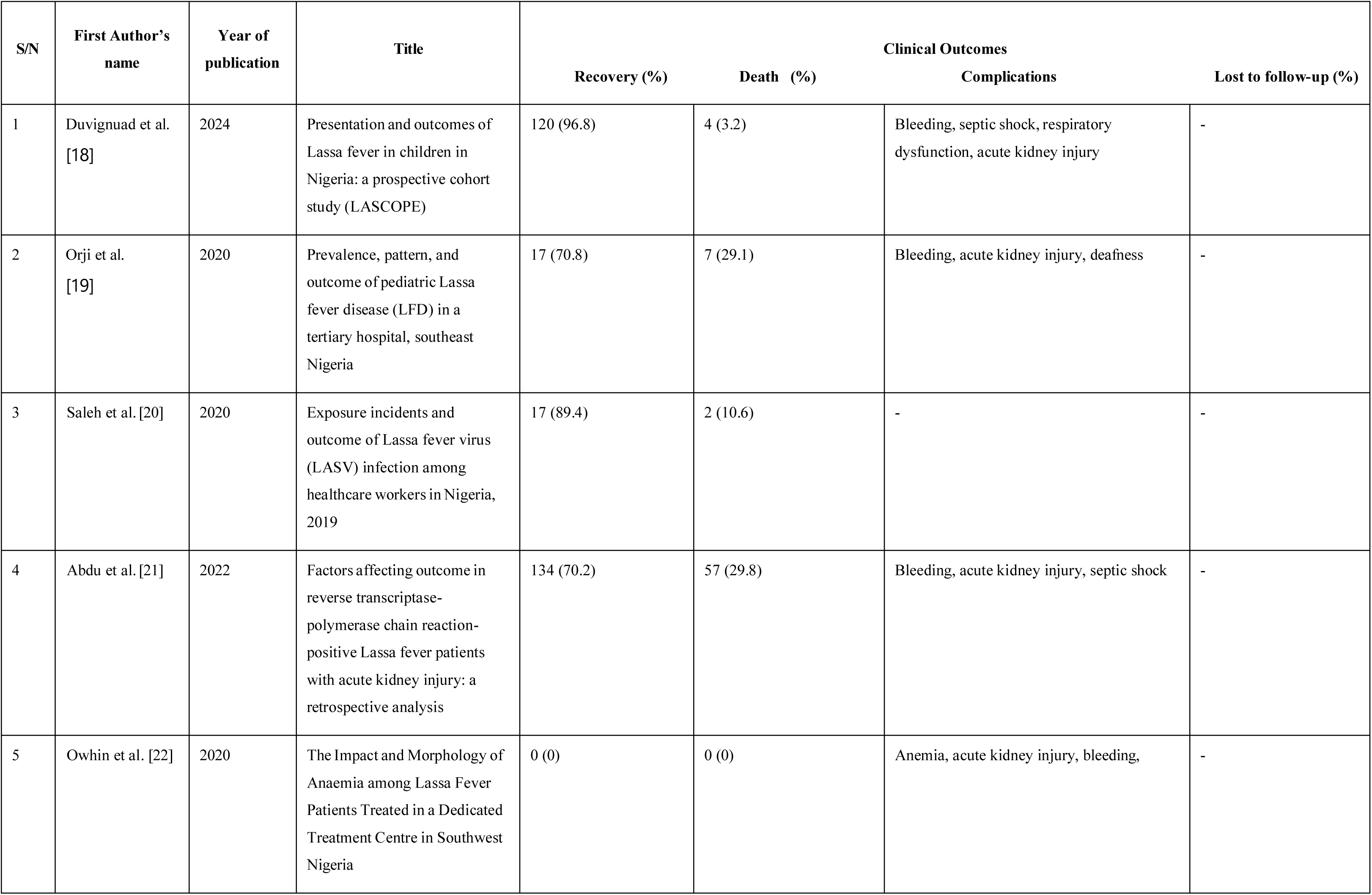
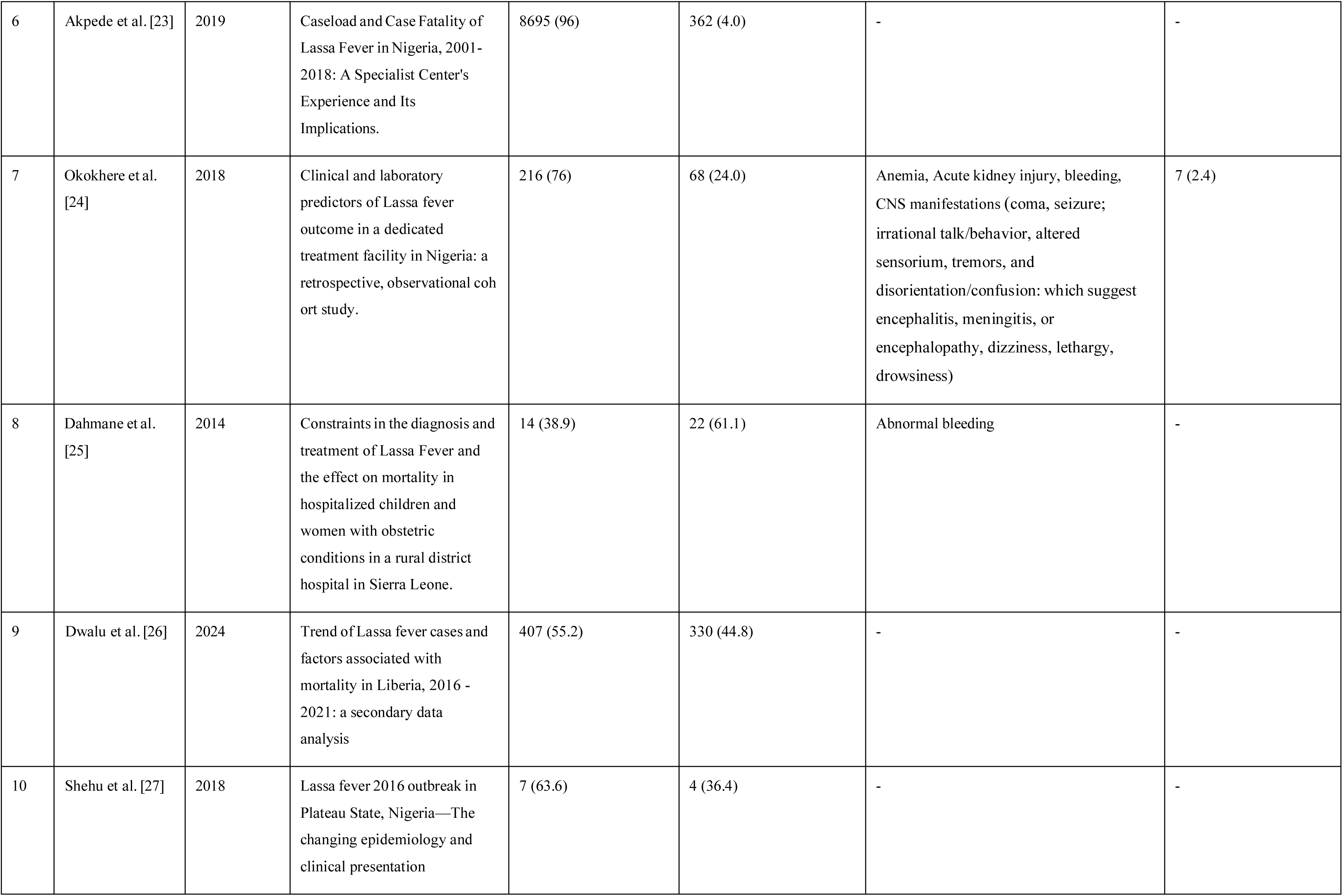
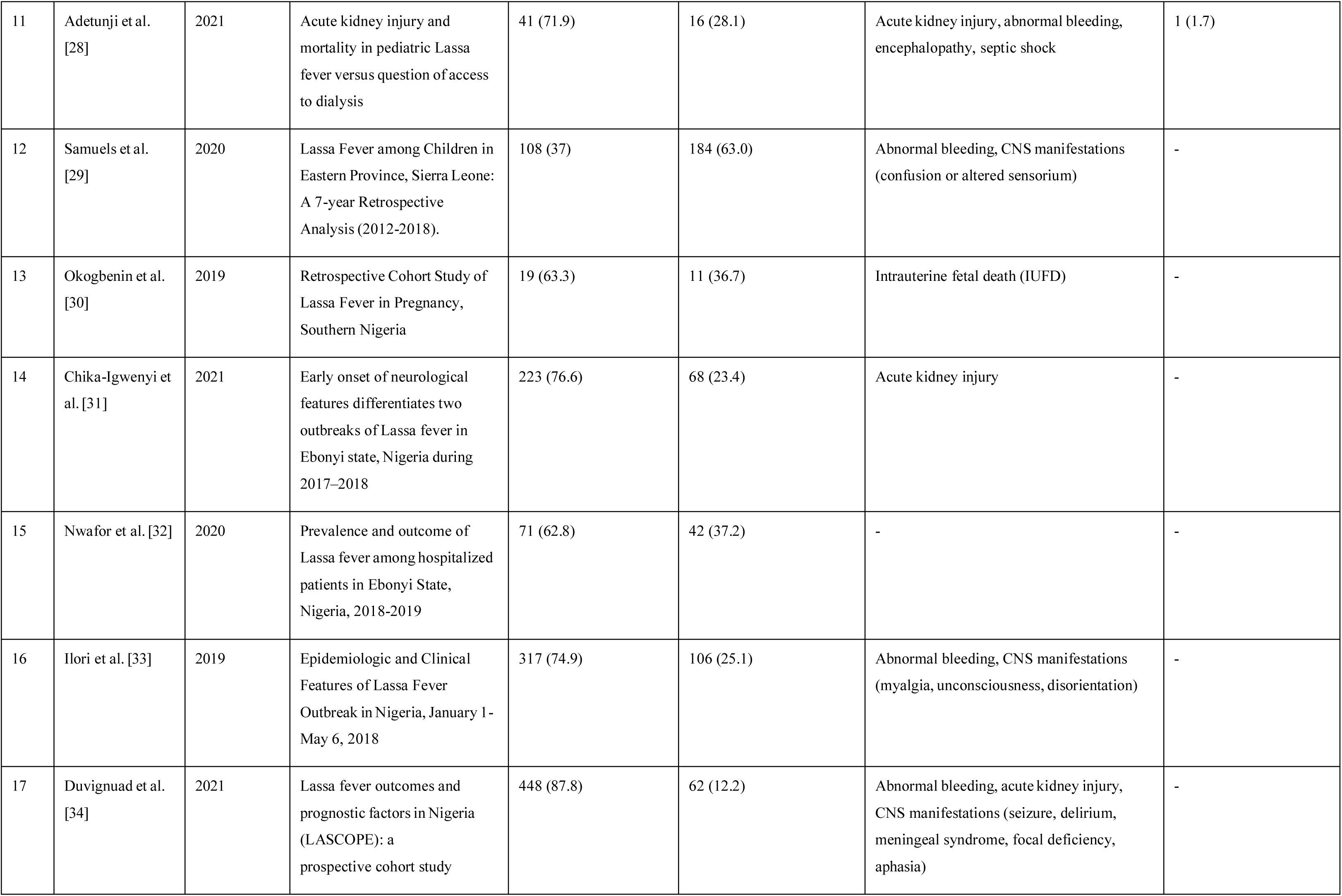
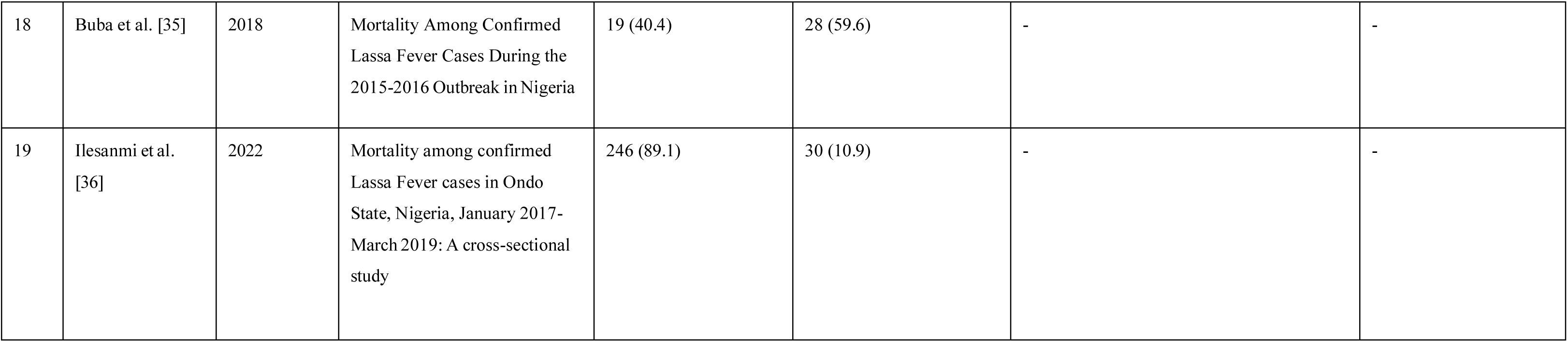
Clinical outcomes of included studies.

Mortality: Mortality rates ranged from 3.2% to 63.0% in the included studies. The mortality rate was highest in the study carried out by Samuels et al. in the Eastern Province of Sierra Leone 2020 (63.0%). [29]

Complications: The majority of the studies included in the review reported complications (12/19; 63.2%). [18–22,24,25,28,34] The most common complications reported included abnormal bleeding (10/19; 52.6%) [18–22,24,25,28,29,33,34] acute kidney injury (8/19; 42.1%) [18,19,21,22,24,28,31,34] and central nervous system (CNS) manifestation (8/19; 42.1%). [24,28,29,33] Sensorineural deafness, another clinically relevant complication was also reported (1/19; 5.3%) in the review. [19] (Figure 4)

Lost to follow-up (LFTU): Only one study reported LFTU (1/19; 5.2%). [24] Okokhere and colleagues reported in 2018 that seven patients were discharged against medical advice (Table 2).

**Figure 4:**
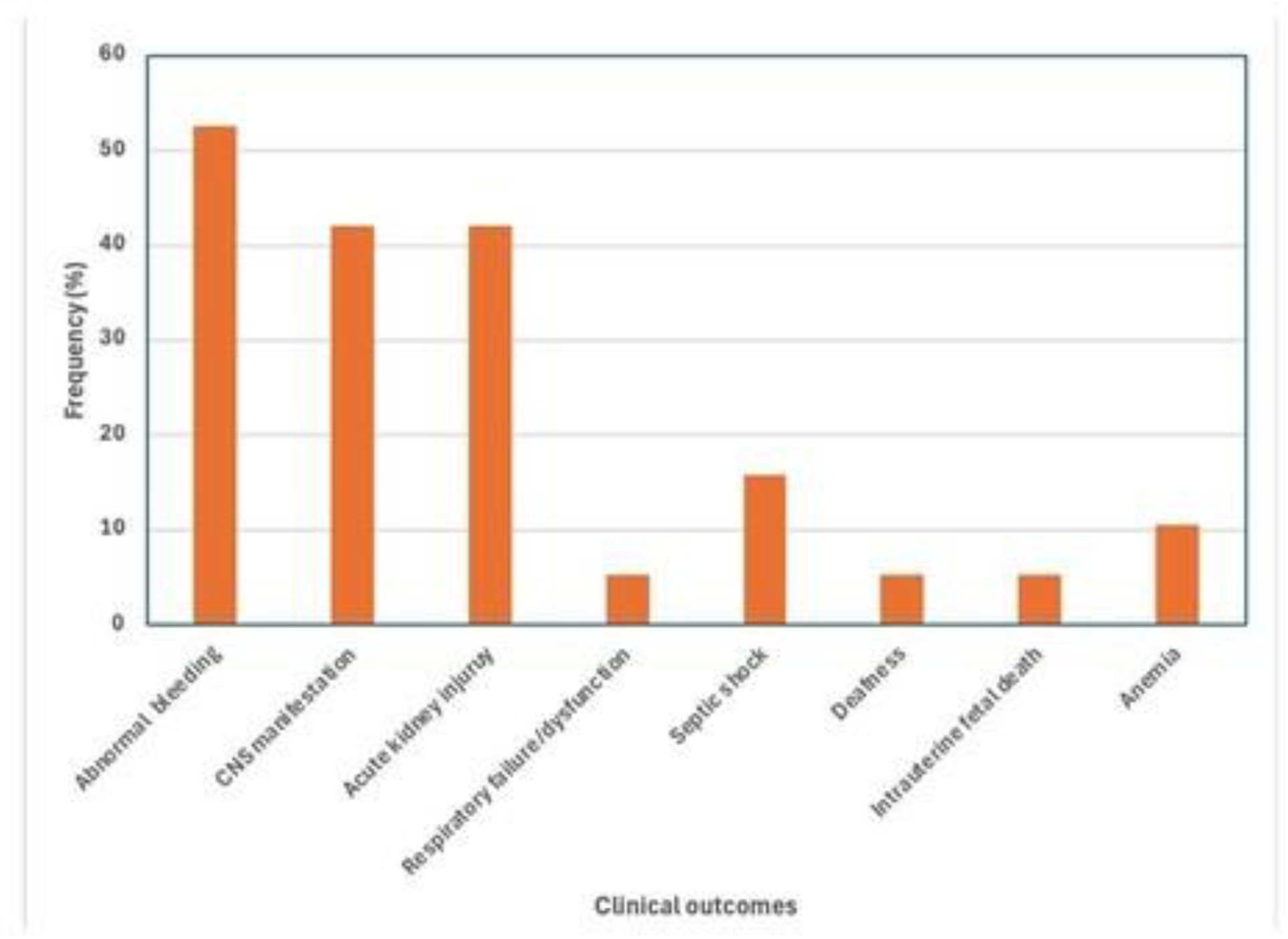
Frequency of complications of Lassa fever.

### Meta-analysis

The pooled proportions of mortality and survival rates among participants with Lassa fever were analyzed based on findings from 18 studies. [17–36] The analysis revealed a pooled mortality rate of 19% (95% CI: 10-32), while the survival rate was found to be 70% (95% CI: 48-86). Notably, there was significant heterogeneity in the outcomes (see Figure 5 and 6).

**Figure 5:**
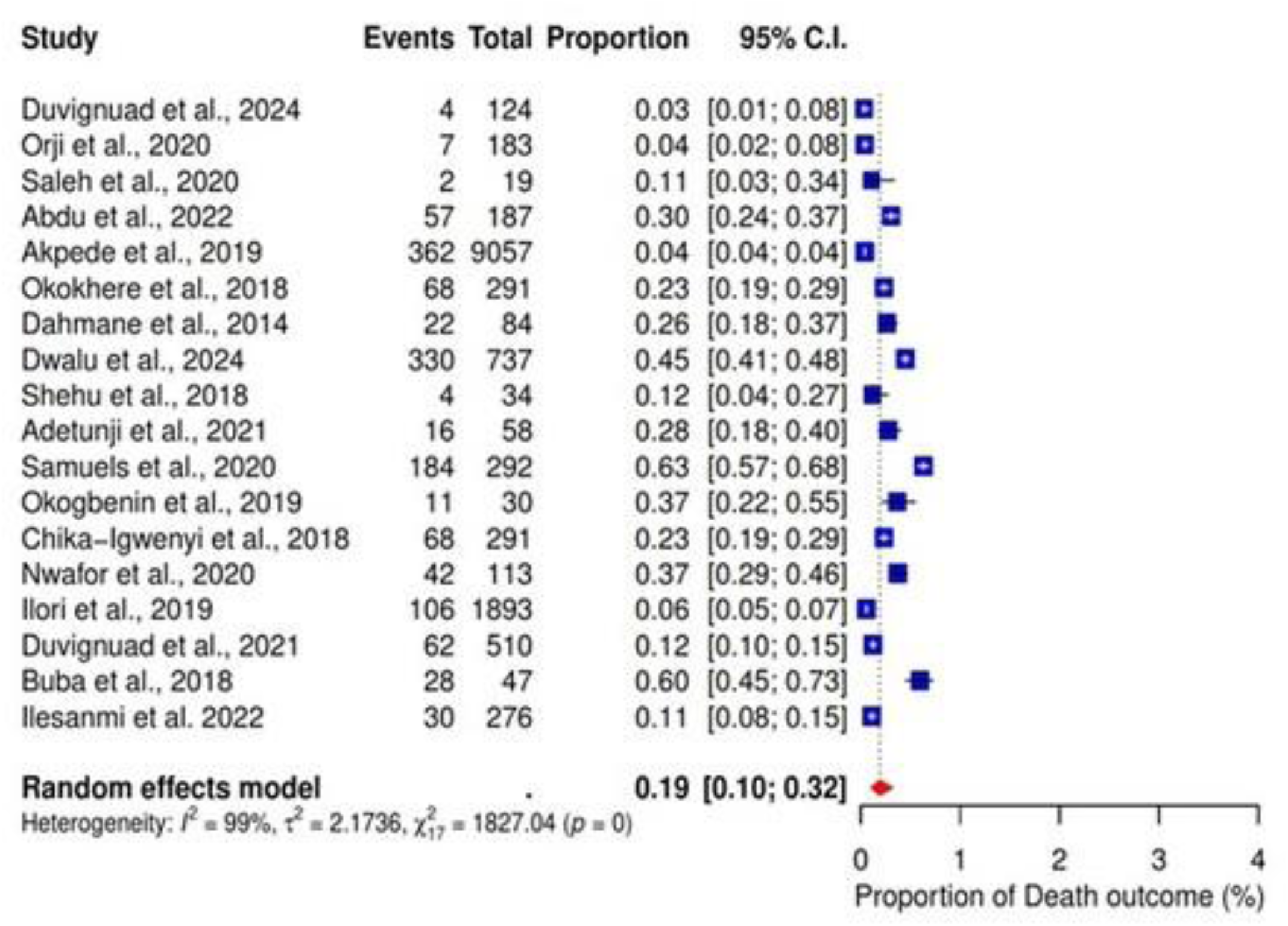
Pooled proportion of death as an outcome of Lassa fever.

**Figure 6:**
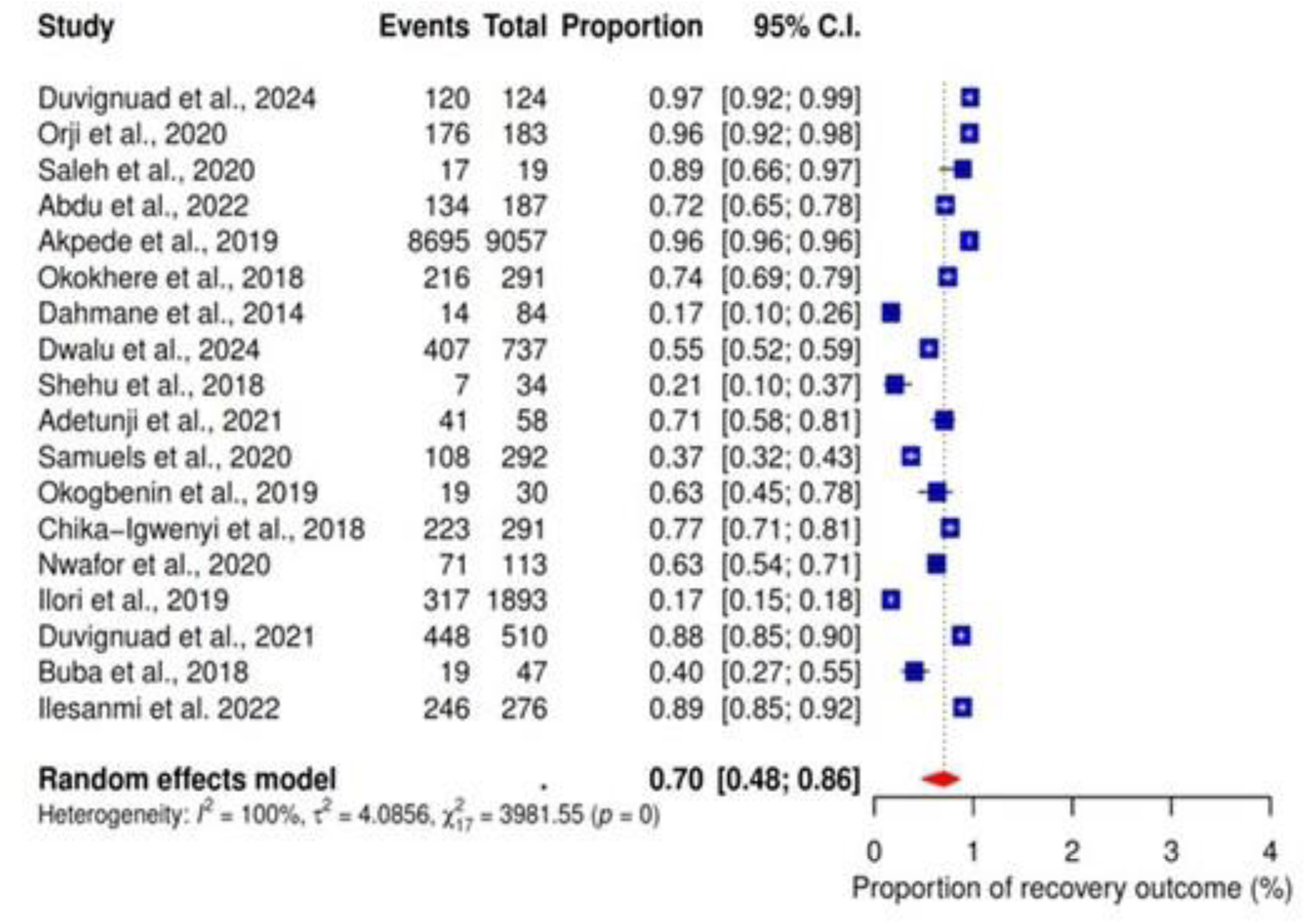
Pooled proportion of recovery as an outcome of Lassa fever.

Subgroup analysis by year of publication showed that the death rate for studies published in years 2018, 2019, 2020, and 2024 were 28% (95% CI: 17-43; I^2^ =90%), 9% (95% CI: 2=05-07; I^2^ =96%), 22% (95% CI: 06-55; I^2^ =97%) and 85% (95% CI: 20-99; I^2^ =97%) (subgroup p value= 0.06) respectively, while the recovery rate of years 2018, 2019, 2020 and 2024 was 55% (95% CI: 35-74; I^2^ =94%), 67% (95% CI: 04-99; I^2^ =100%), 78% (95% CI: 45-94; I^2^ =97%) and 85 (95% CI: 20-99; I^2^ =97%) respectively (subgroup p value<0.01) (Figures 5 and 6).

Group-based analysis among the studies conducted in Nigeria showed a death rate of 13% (95% CI: 06-23; I^2^ =98%) in Owo and 6% (95% CI: 05-07; I^2^ =98%) in Abakaliki (subgroup p value<0.01), while recovery rates were 83% (95% CI: 64-93; I^2^ =99%) in Owo and 55% (95% CI: 03-98; I^2^ =98%) respectively (subgroup p value<0.01). (Figure 7-10)

**Figure 7:**
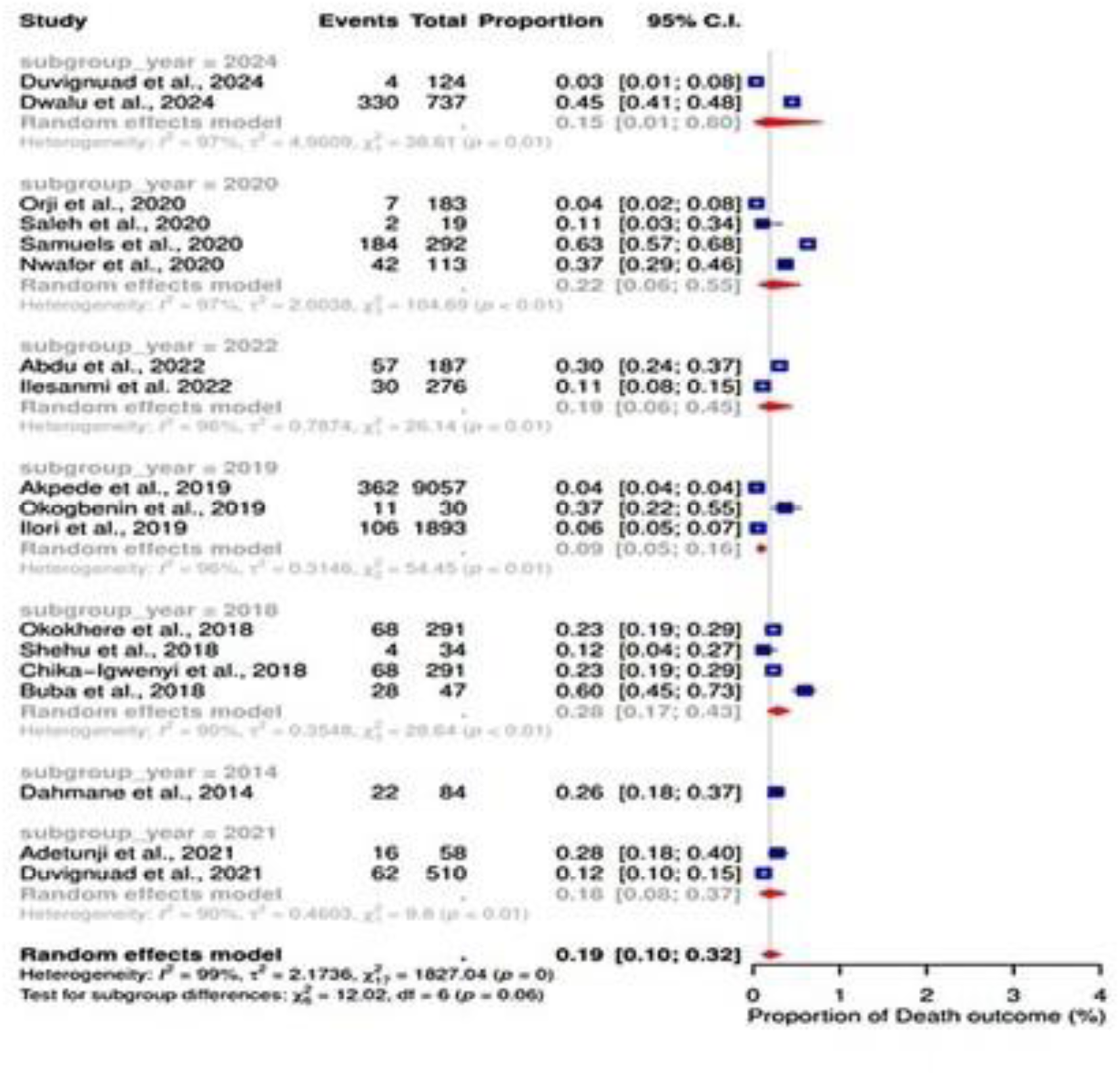
Subgroup results of death outcome by year of publication.

**Figure 8:**
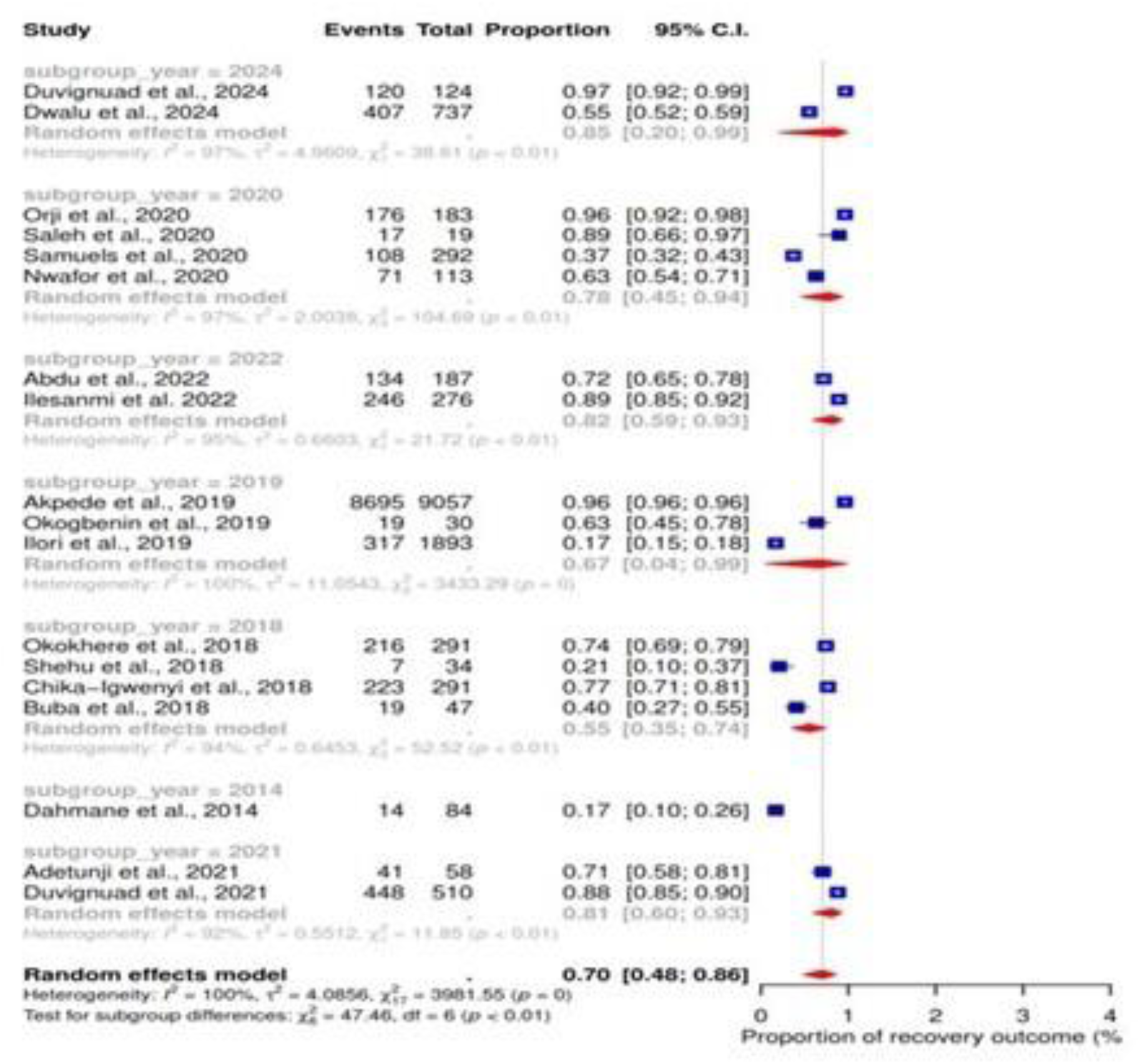
Subgroup analysis of survival outcome by year of publication.

**Figure 9:**
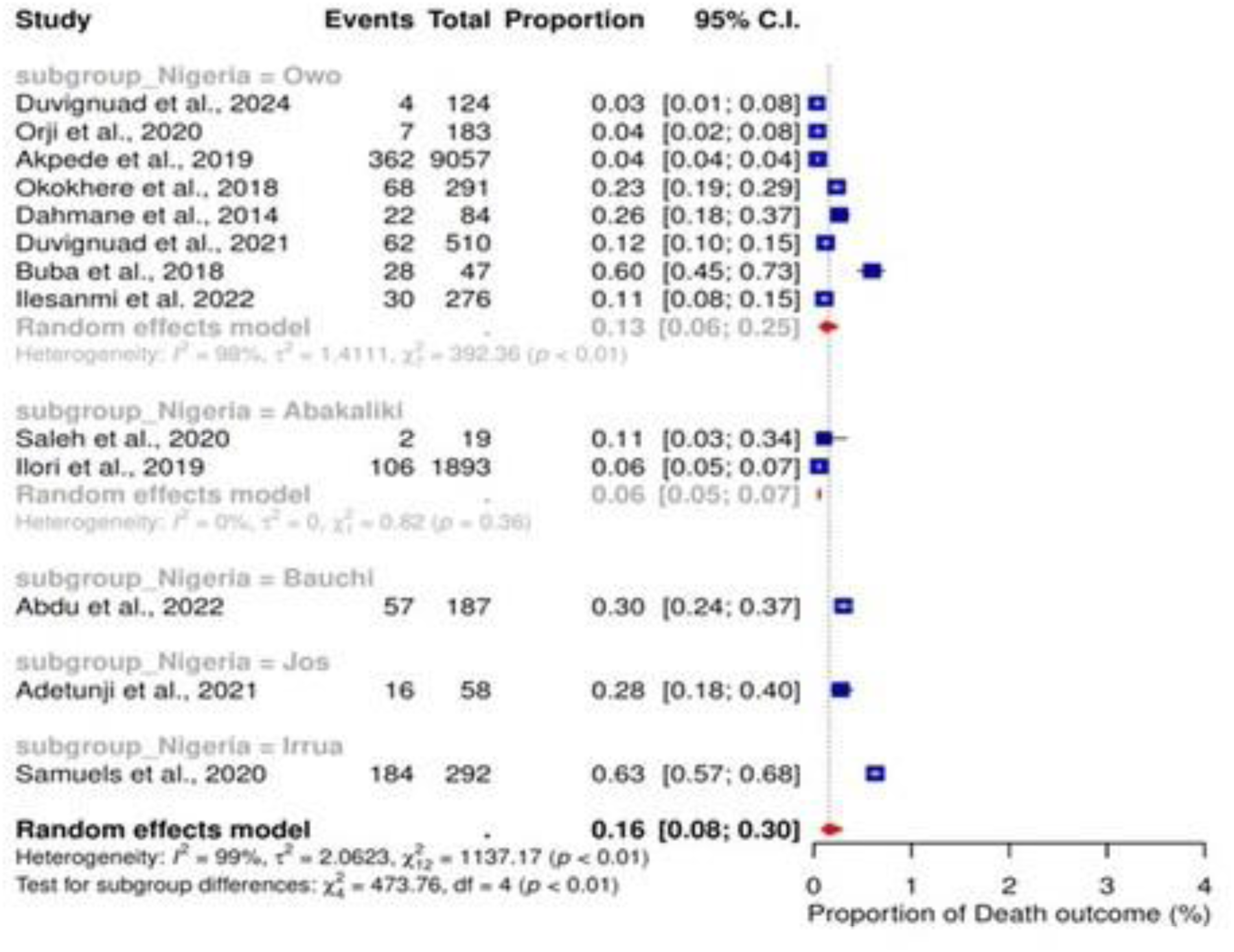
Subgroup analysis of mortality outcome by locations in Nigeria.

**Figure 10:**
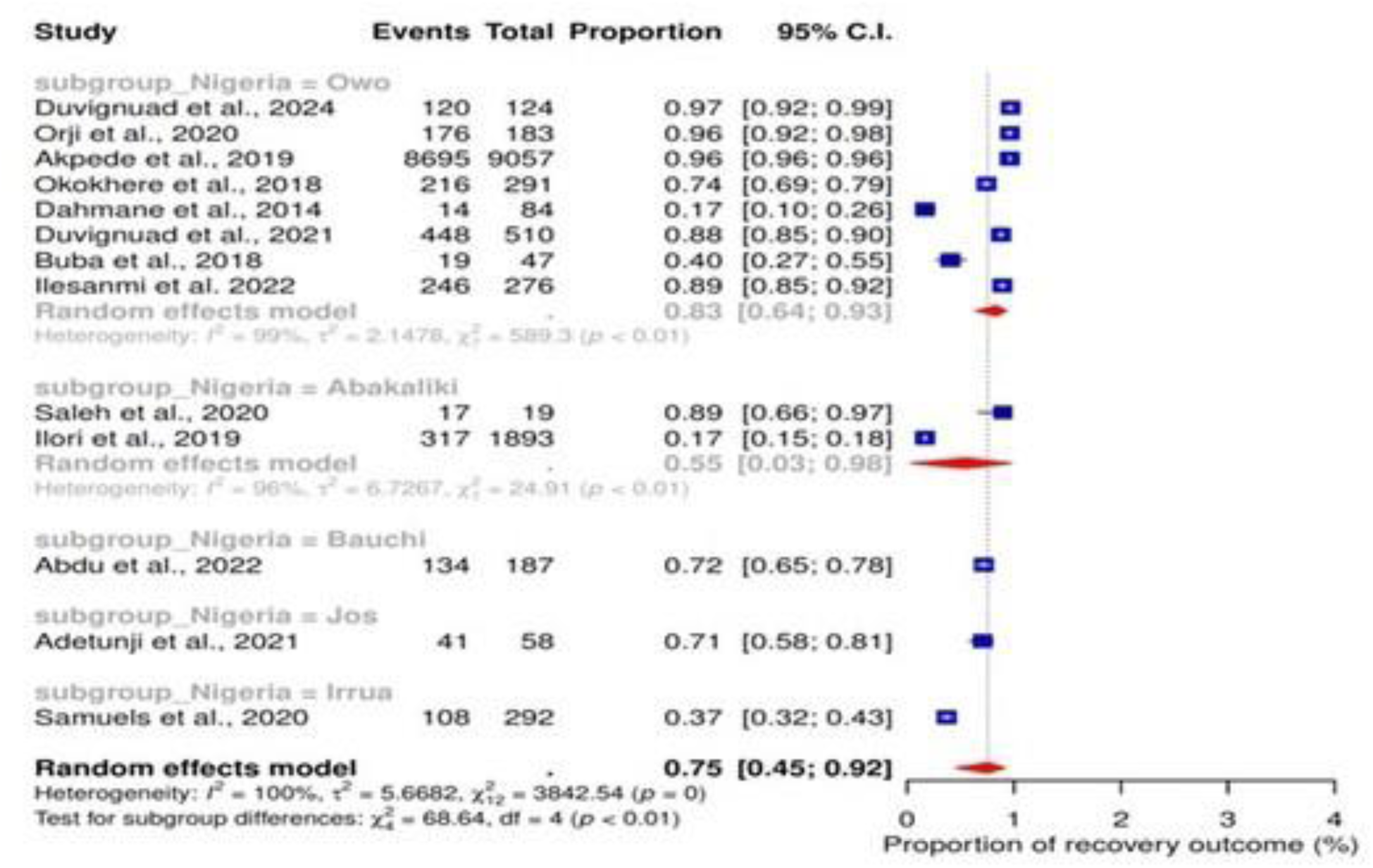
Subgroup analysis of survival outcome by locations in Nigeria.

### Abnormal bleeding

We conducted a meta-analysis across 10 studies that reported abnormal bleeding as a clinical outcome of Lassa fever infection. The finding showed a pooled proportion of 17% (95% CI; 9-30; I^2^=98%) (figure 8). Furthermore, the sub-group analysis result showed a pooled proportion of 22% (95% CI; 14-34; I^2^=93%) (subgroup p-value=<0.01) of abnormal bleeding across studies conducted in Owo (figure 11 and 12).

**Figure 11:**
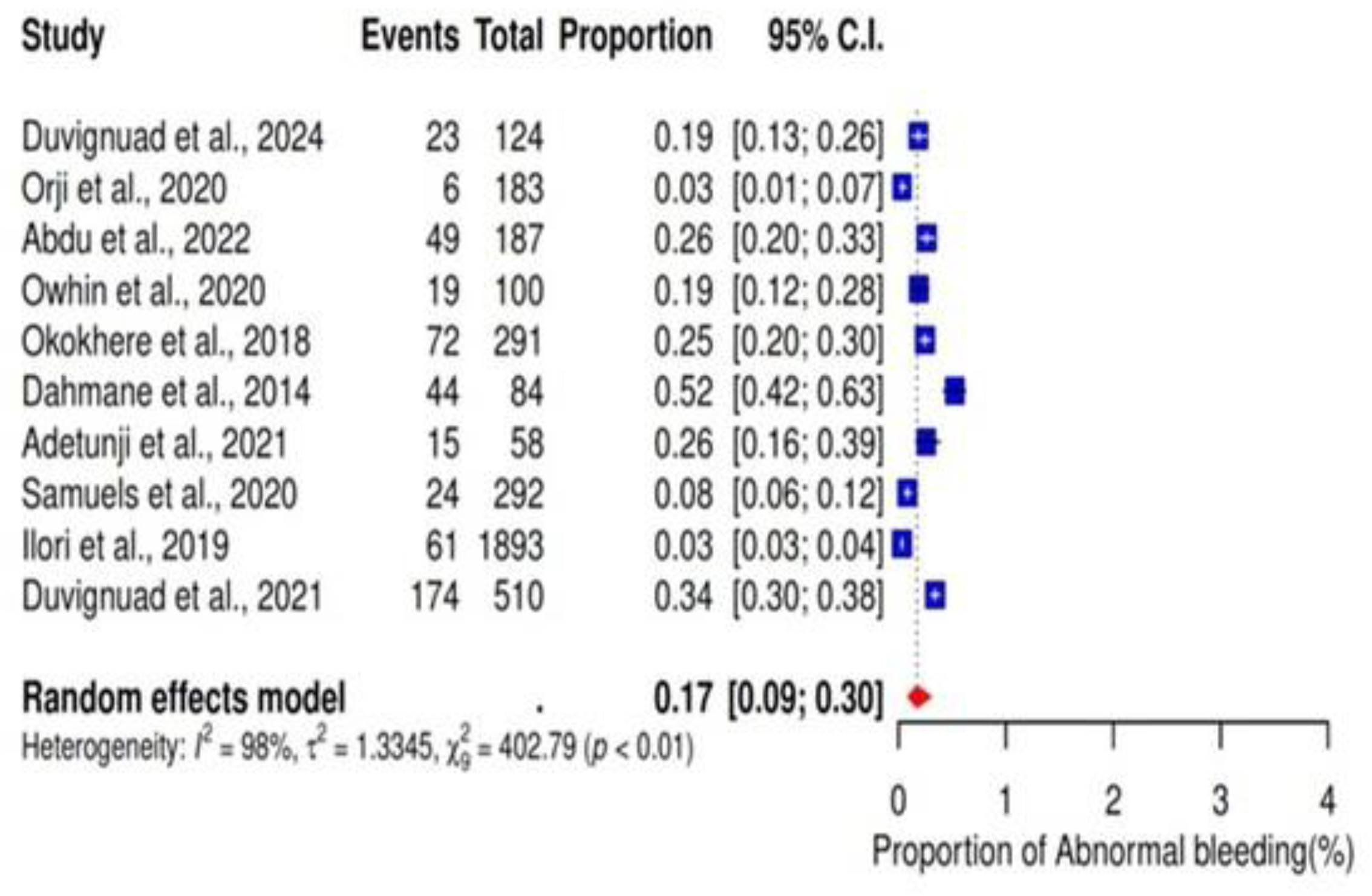
Forest plot showing pooled proportion of abnormal bleeding as a clinical outcome of Lassa fever.

**Figure 12:**
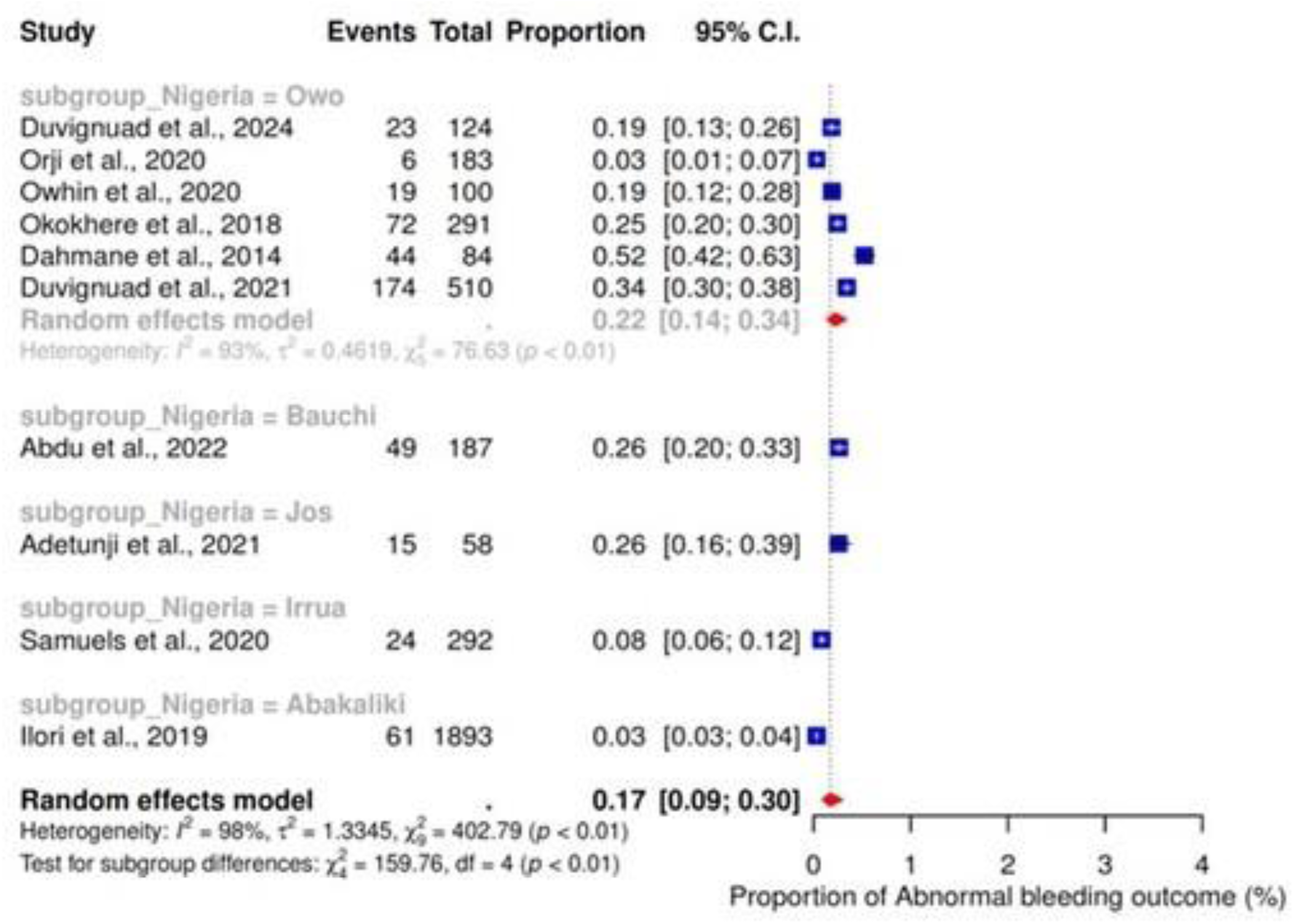
Subgroup analysis of abnormal bleeding by locations in Nigeria.

### Acute kidney injury outcome

Acute kidney injury was reported across 7 studies as a clinical outcome for Lassa fever infection. Our findings showed a pooled proportion of 19% (95% CI; 13-26; I^2^=89%) (figure 10). Based on group analysis among studies conducted in Nigeria, the pooled proportion of 19% (95% CI; 12-28; I^2^=91%) was found in Owo (subgroup p value=0.03) (Figure 13).

**Figure 13:**
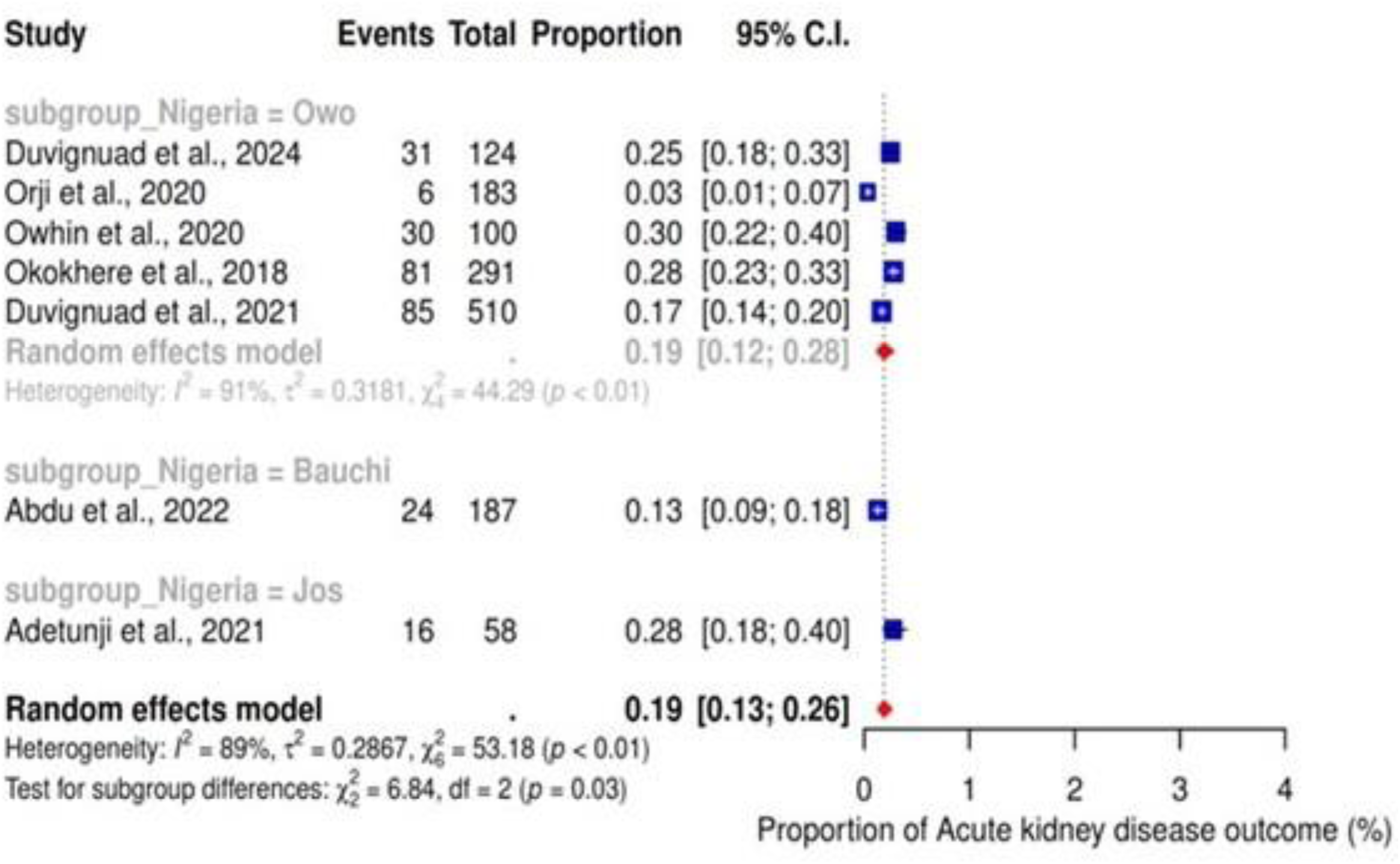
Subgroup analysis of Acute kidney injury by locations in Nigeria.

### CNS manifestation

We conducted a meta-analysis across 5 studies that reported CNS manifestations as a clinical outcome of Lassa fever infection in their studies. The finding showed a pooled proportion of 15% (95% CI; 6-32; I^2^=98%) (figure 14).

**Figure 14:**
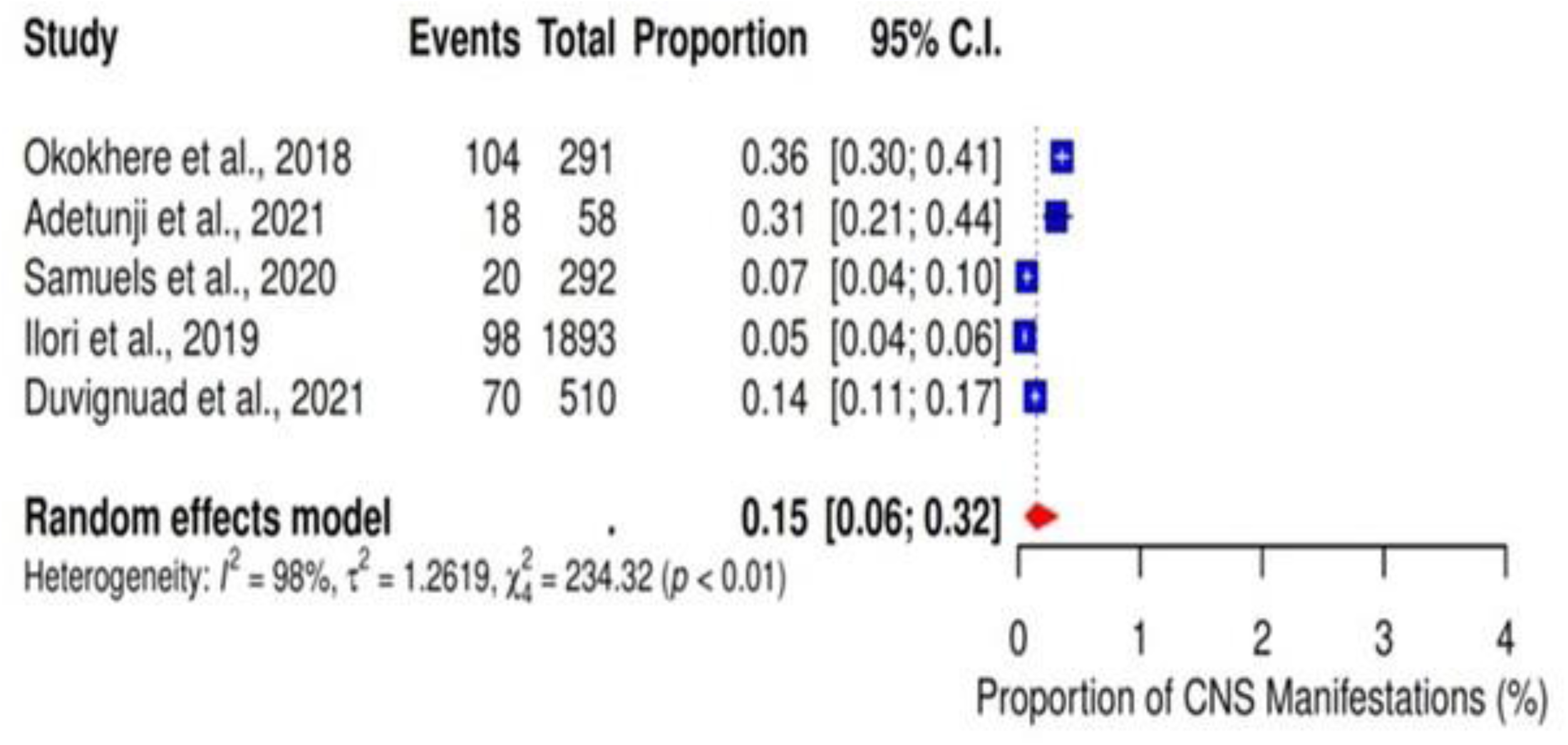
Pooled proportions of CNS manifestations as a clinical outcome of Lassa fever.

### Publication bias and sensitivity analysis

The funnel plots revealed significant asymmetry (figures 15 to 19). To further evaluate the impact of individual studies on the pooled proportion, a leave-one-out analysis was performed. The results indicated only minor variations from the original summary proportion. This suggests that the conclusions derived from this meta-analysis should be interpreted with caution.

**Figure 15:**
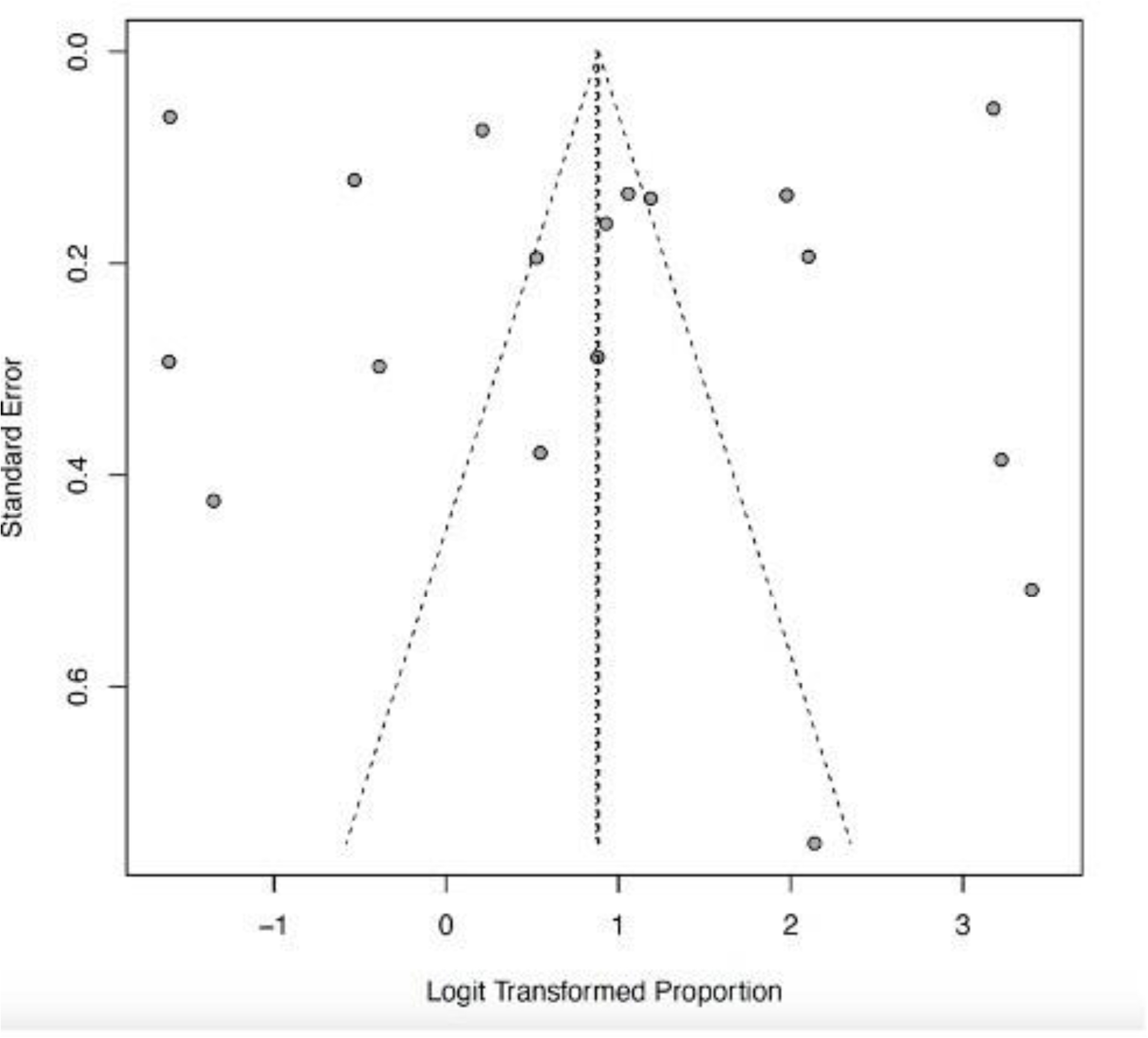
Funnel plot of recovery as a clinical outcome of Lassa fever.

**Figure 16:**
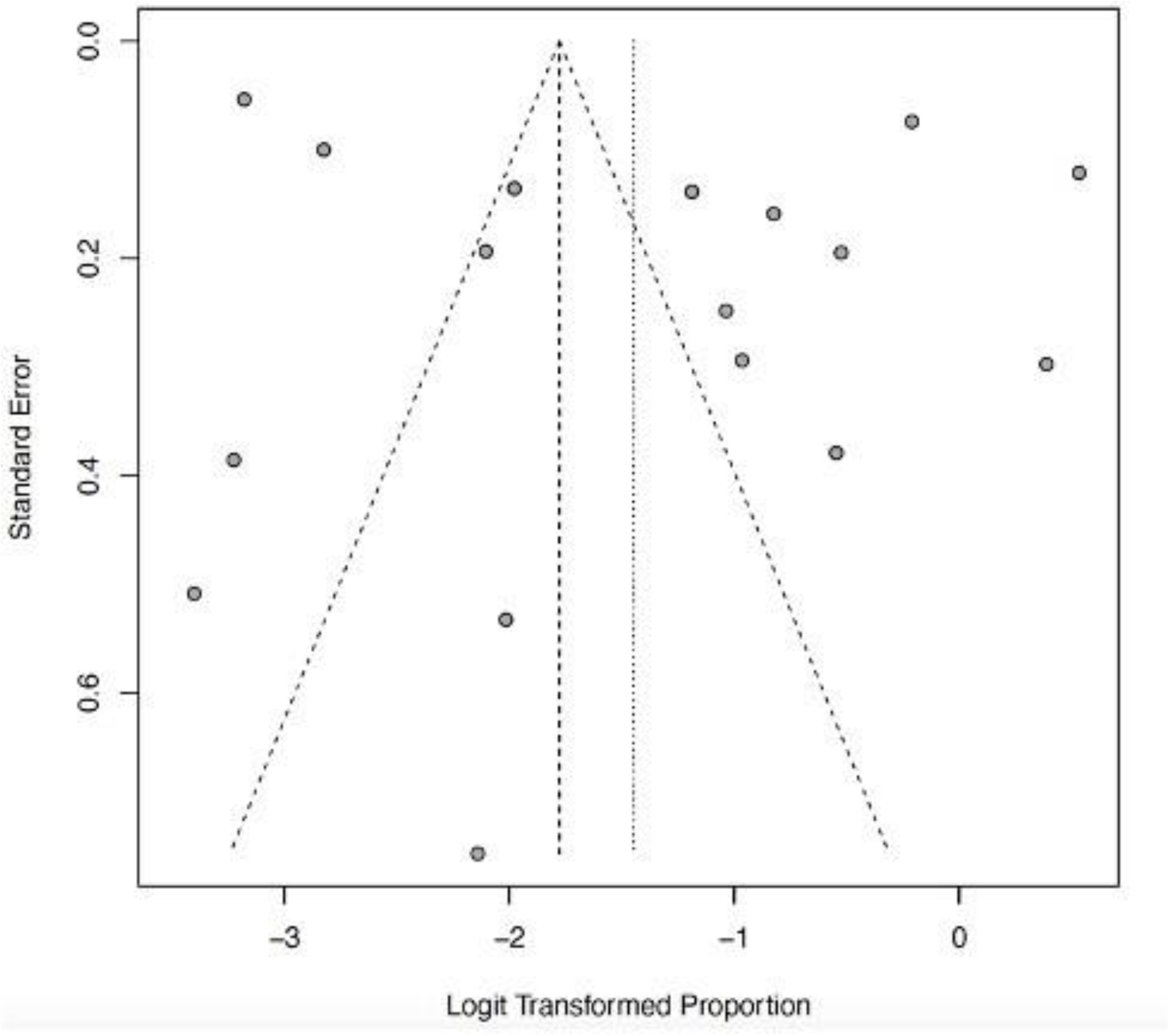
Funnel plot of death as a clinical outcome of Lassa fever.

**Figure 17:**
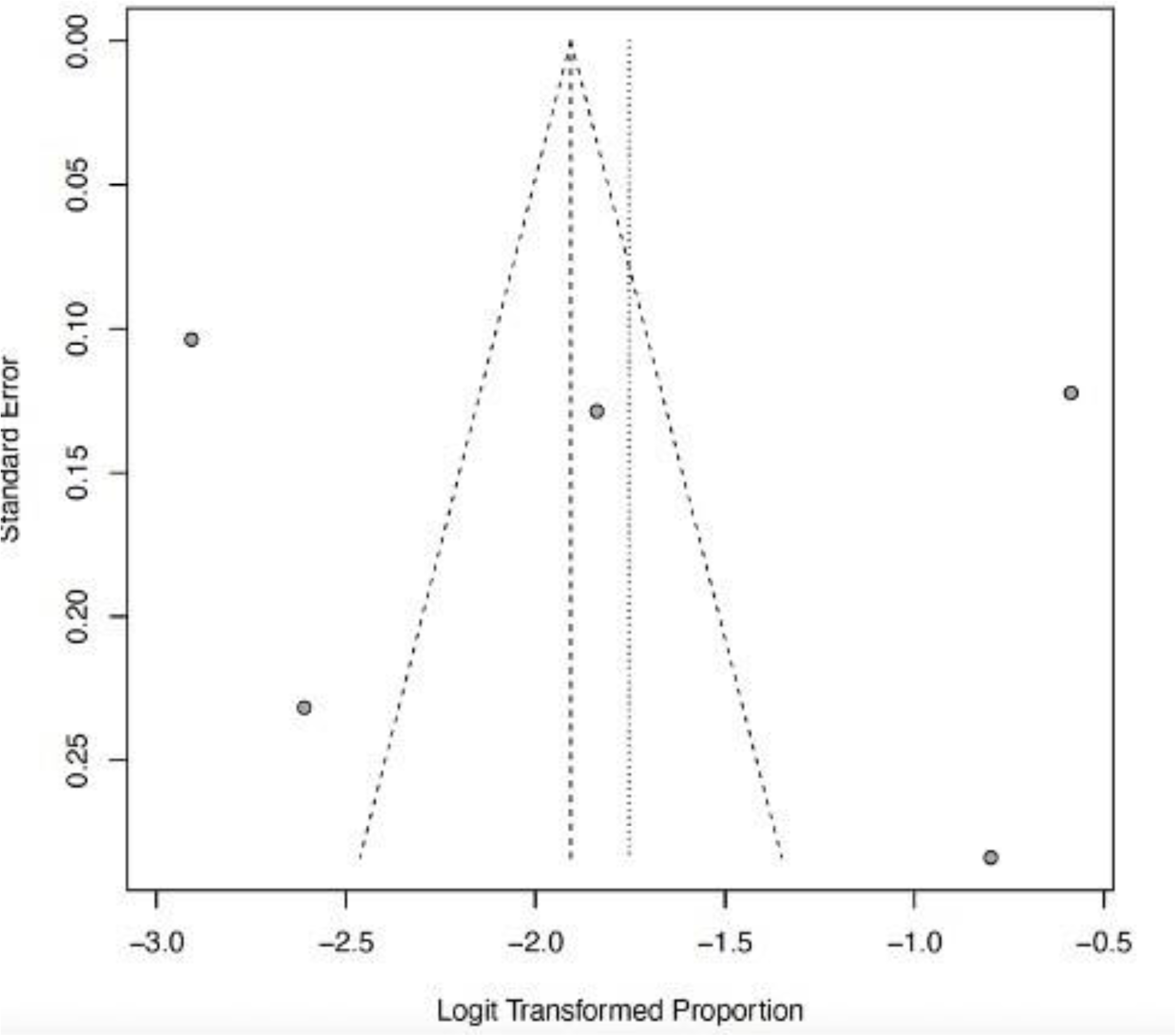
Funnel plot of CNS manifestations as a clinical outcome of Lassa fever.

**Figure 18:**
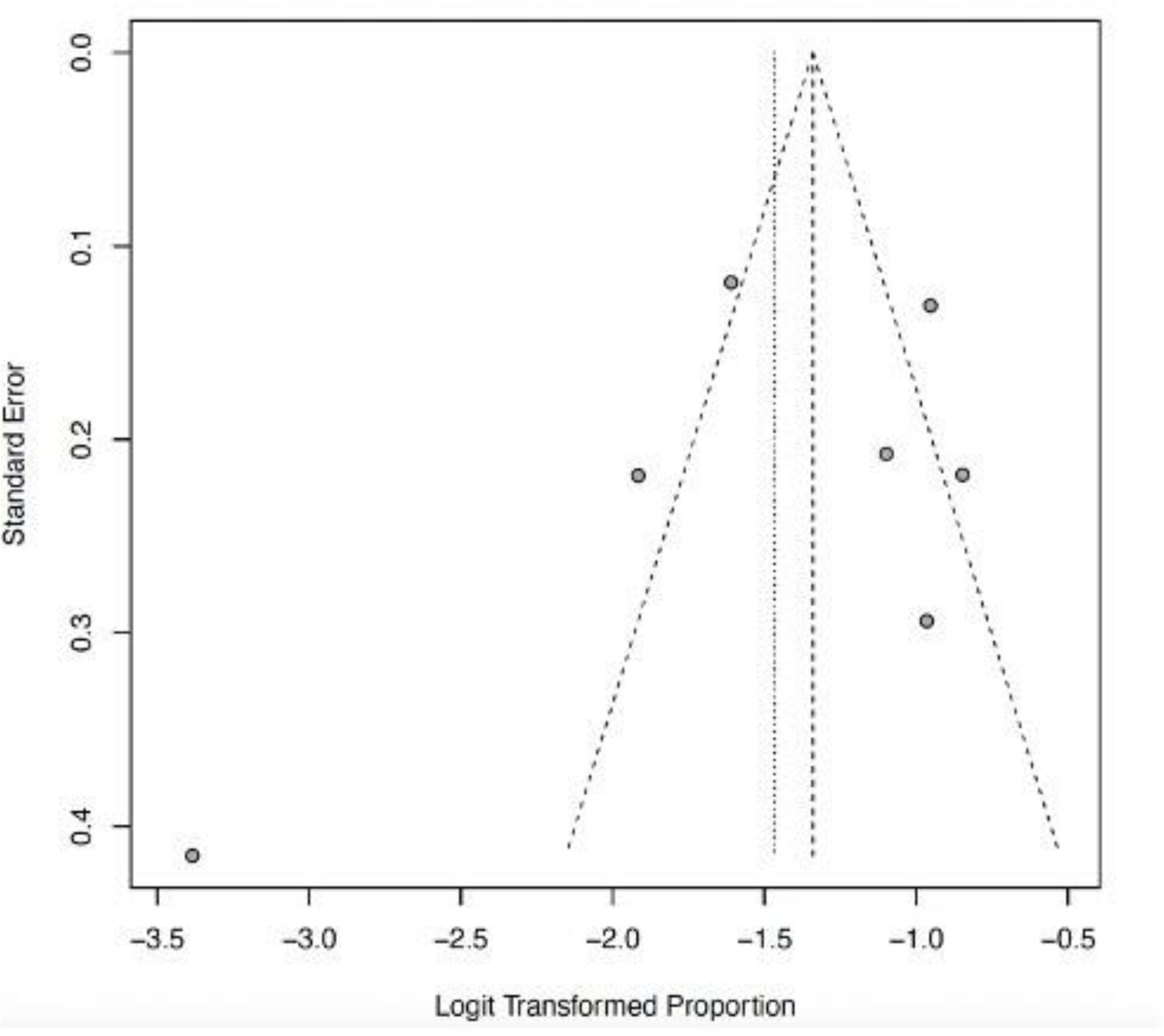
Funnel plot of acute kidney disease as a clinical outcome of Lassa fever.

**Figure 19:**
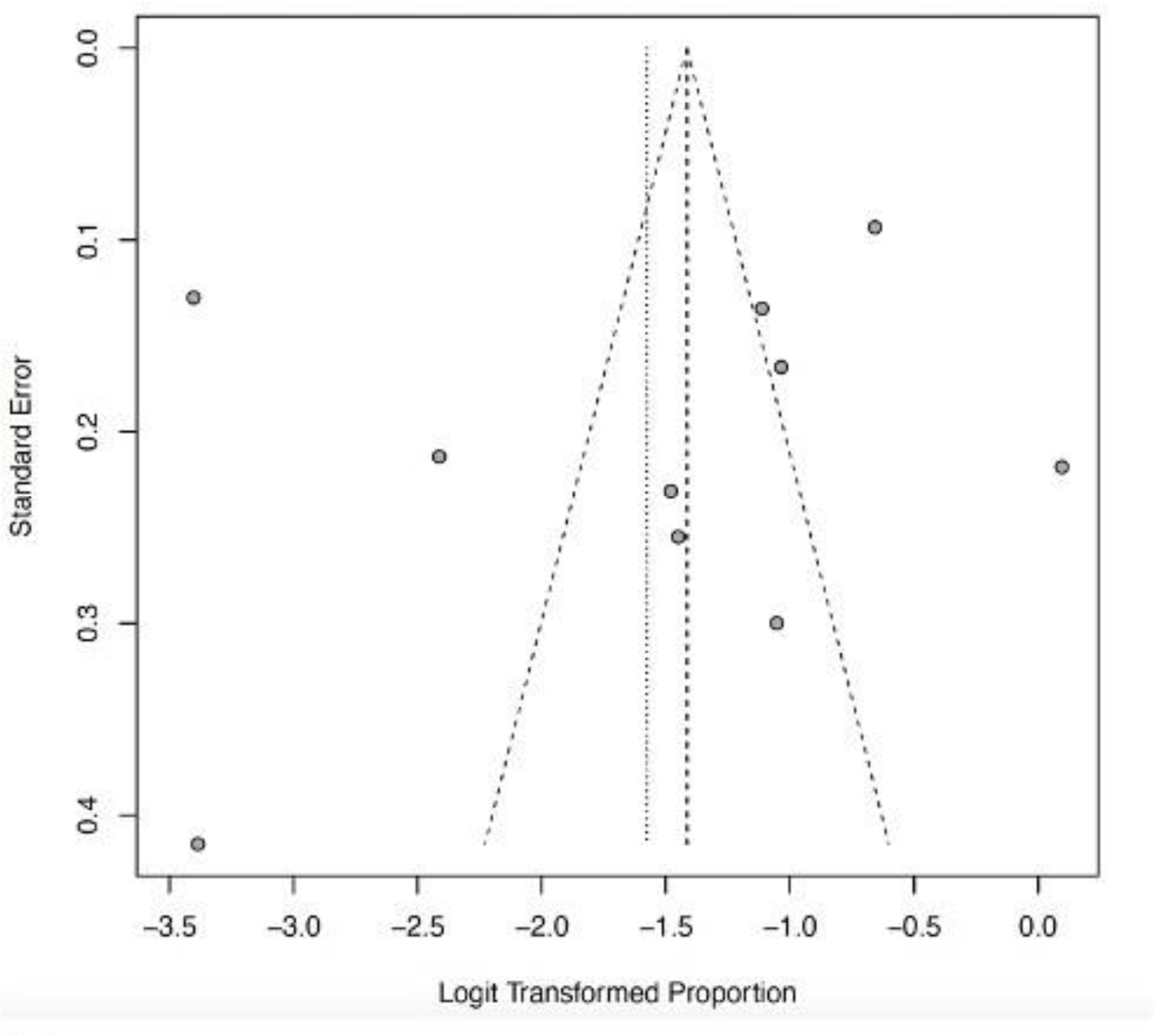
Funnel plot of abnormal bleeding as a clinical outcome of Lassa fever.

## Discussion

This systematic review and meta-analysis (SRMA) provide a comprehensive overview of the clinical outcomes of LF in West Africa. Our study revealed a pooled mortality rate of 19% (95% CI: 10-32) and common complications such as acute kidney injury, abnormal bleeding, and CNS manifestations. By synthesizing data from 19 studies involving 4177 patients across 3 high-burden countries, our study highlights the complexity of clinical outcomes of LF in the West African sub-region with significant implications for public health policy and clinical practice.

We reported studies predominantly from Nigeria (88.9%) in keeping with data that reported Nigeria as the country with the highest burden of disease in the region. [37] In Nigeria, most of the included studies were concentrated in the three LF hotspots (Owo, Irrua, and Abakaliki). The tropical climate and high temperature in Nigeria favour the multiplication and survival of the LF vector rodent (*Mastomys natalensis)* leading to increased transmission and virulence of the virus during disease epidemics amongst overcrowded rural populations. [7] This is further corroborated by evidence showing that LASV is hosted by other rodent species: the African wood mouse (*Hylomyscus pamfi)* and the Guinean multimammate mouse (*Mastomys erythroleucus)* identified in Nigeria. [6] Our study design was predominantly retrospective cohort studies (89.5%). The Lassa fever clinical course and prognostic factors in an epidemic context (LASCOPE) conducted by Duvignaud and colleagues in Owo in 2019 is the largest prospective cohort study ever conducted on Lassa fever [34] in Nigeria following the inclusion of LF in the blueprint priority list of diseases for intensified research and development (R & D) requiring urgent medical countermeasures in 2019 by the WHO. [9] Although ribavirin is used for the treatment of LF, it has yet to secure regulatory approval by the WHO as no well-designed randomized control trial (RCT) has been conducted on LF treatment. [38] Most of the included studies were published within the last decade (2014-2024) highlighting the fact that LF was neglected for about 50 years since its discovery in 1969. The disease gained the WHO’s attention in 2019 following an unusual increase of 633 laboratory-confirmed LF cases in Nigeria in 2018. Currently, efforts are being made to accelerate medical countermeasures such as rapid diagnostic tests, vaccines, and therapeutics to improve detection and prompt management of cases. [39,40] Males were more affected than females in the included studies. This may be because males are more mobile and are likely to engage in high-risk occupations (e.g., farming, hunting) that increase exposure to infected rodents in rural areas. [20] Furthermore, males generally tend to have poor healthcare-seeking behaviour leading to poor outcomes. Our review equally reported the occurrence of a high percentage of included studies (94.7%) in rural settings. Lassa fever infection commonly occurs in rural endemic areas where there is an abundance of rodent reservoirs (*mastomys natalensis*) and a narrow proximity between humans and their environment.

The pooled high mortality rate of LF of 19% (95% CI: 10-32) reported in our study highlights the considerable threat LF poses to regional public health security. This may be attributed to a low level of awareness of the disease among the public and healthcare workers often leading to missed or delayed diagnosis and treatment, particularly in the rural endemic areas. This is further worsened by the absence of readily available point-of-care assays for early diagnosis coupled with the long turnaround time due to transport and other logistic issues associated with reliance on a PCR-based diagnosis. Lassa fever symptoms are highly variable with about 80% of cases presenting with no or mild symptoms similar to other tropical febrile illnesses such as malaria, and typhoid fever. For instance, two retrospective studies conducted in Sierra Leone by Dahmane et al. [25] and Samuels et al. [29] reported high mortality rates of 61% and 63% respectively. This may be attributed to the high virulence of the circulating LASV lineage IV found in Liberia and Sierra Leone [40] and delayed access to medical care and treatment due to weak health infrastructure. Furthermore, certain high-risk groups are more susceptible to severe disease and death. Pregnant women, especially in the third trimester, are at increased risk of maternal and fetal mortality. A study done by Okogbenin and colleagues in 2019 reported a mortality rate of 36.7% among pregnant cohorts. [30] This has been attributed to the increased replication of the virus within the rapidly dividing/highly vascularized placental tissue. Also, the similarity of Lassa fever symptoms such as nausea, headache, and abdominal pain with early pregnancy symptoms may further delay diagnosis by an unsuspecting clinician leading to poor outcomes when the infection is severe. [30] The most common complications of LF reported in this review are acute kidney injury (AKI) at a pooled proportion of 19% (95% CI; 13-26; I^2^=89%), followed by abnormal bleeding at a pooled proportion of 17% (95% CI; 9-30; I^2^=98%), and CNS manifestations at a pooled proportion of 15% (95% CI; 6-32; I^2^=98%). While abnormal bleeding is a poor surrogate for Lassa fever diagnosis, its presence in a patient with acute febrile illness should increase the suspicion of LF by the clinician. The underlying mechanism of abnormal bleeding in LF remains poorly understood but has been attributed to endothelial barrier disruption, abnormal coagulation, and dysfunctional platelet aggregation. [41] Other complications of LF such as AKI and CNS involvement reflect minimal to extensive inflammation due to direct viral cytopathy and other pathogenetic mechanisms such as cytokine storm, oxidative stress, and endothelial damage. [41] Neurological complications are rare at the early stage of the disease but may be seen in late stages, in severely ill patients with high CFR. [42] Sensorineural deafness, another important but relatively uncommon complication, has been attributed to viral-induced immunological injury to the structures of the inner ear and is said to occur in 25% of cases. An animal study (murine model) has shown that LF causes damage to the cochlear hair cells as well as degeneration of the spiral ganglion cells of the auditory nerve. [43]

Our study had a few limitations. We only screened studies published in the English Language between 2004 to 2024 and within the West African region. This time and language restriction could impact the generalizability of our study findings. Furthermore, there was significant variability between the studies such as differences in study design, population characteristics, or healthcare settings which could affect the pooled outcome and may potentially impact the findings of the study.

## Conclusions

In conclusion, our findings suggest that one out of every five patients hospitalized for Lassa fever is likely to die in West Africa. The high pooled mortality rate and severe complications reported in this study underscore the need for risk communication and community engagement as well as (re)training of healthcare workers to improve early diagnosis and case management of LF. Further research is needed to urgently accelerate the development of medical countermeasures such as effective vaccines, potent antiviral therapies, and novel diagnostic assays to address this emerging viral threat.

## Data Availability

All data produced in the present work are contained in the manuscript.

https://www.medRxiv.com

## Acknowledgments

The authors appreciate the support given by the Nigerian Institute for Medical Research Foundation [Grant Number NF-GMTP-24-152809] for the study.

## Supporting information

S1 Text. Search strategy

S2 Text. PRISMA checklist

S3 Text. Data extraction sheet

**Supplementary File 1:**
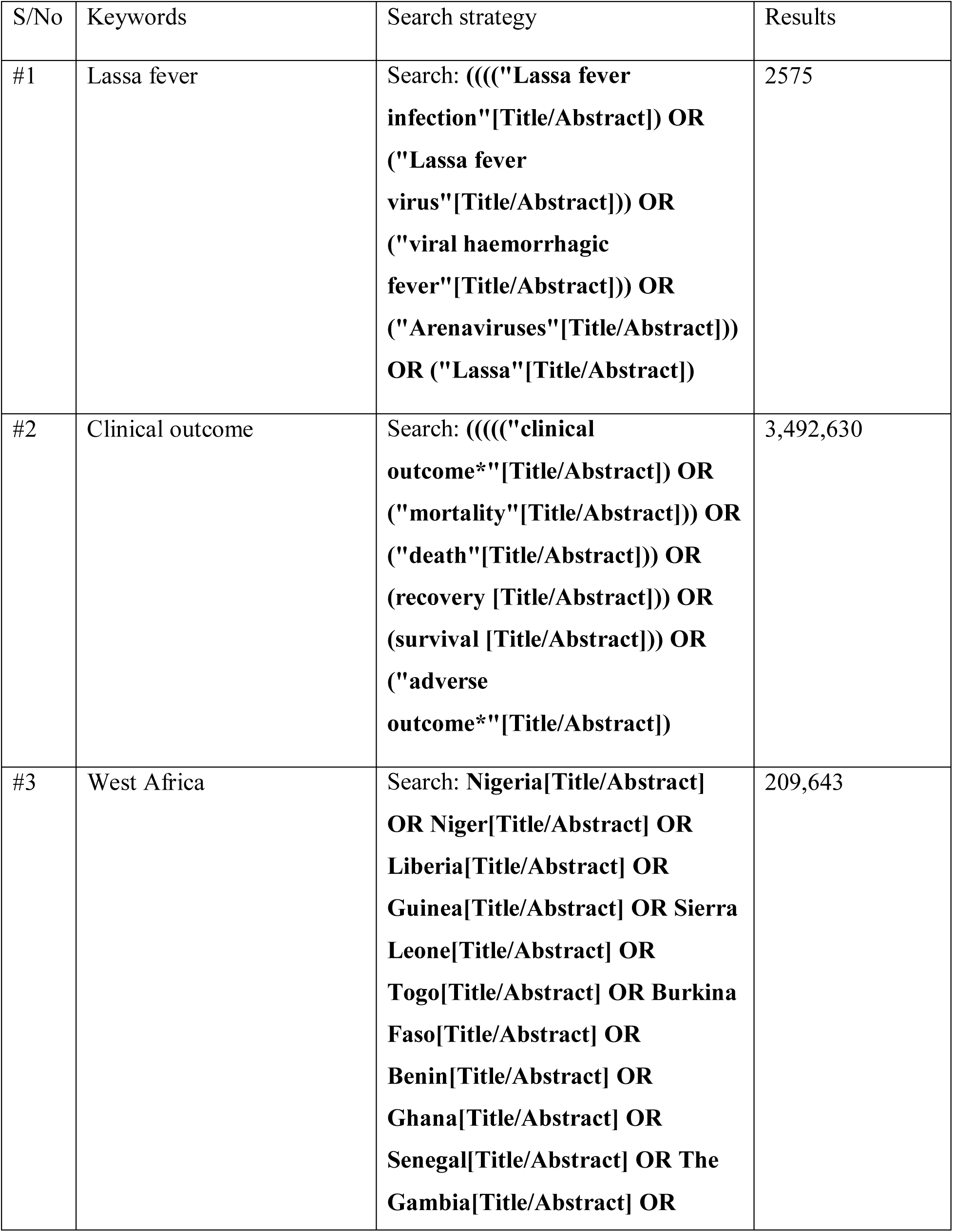
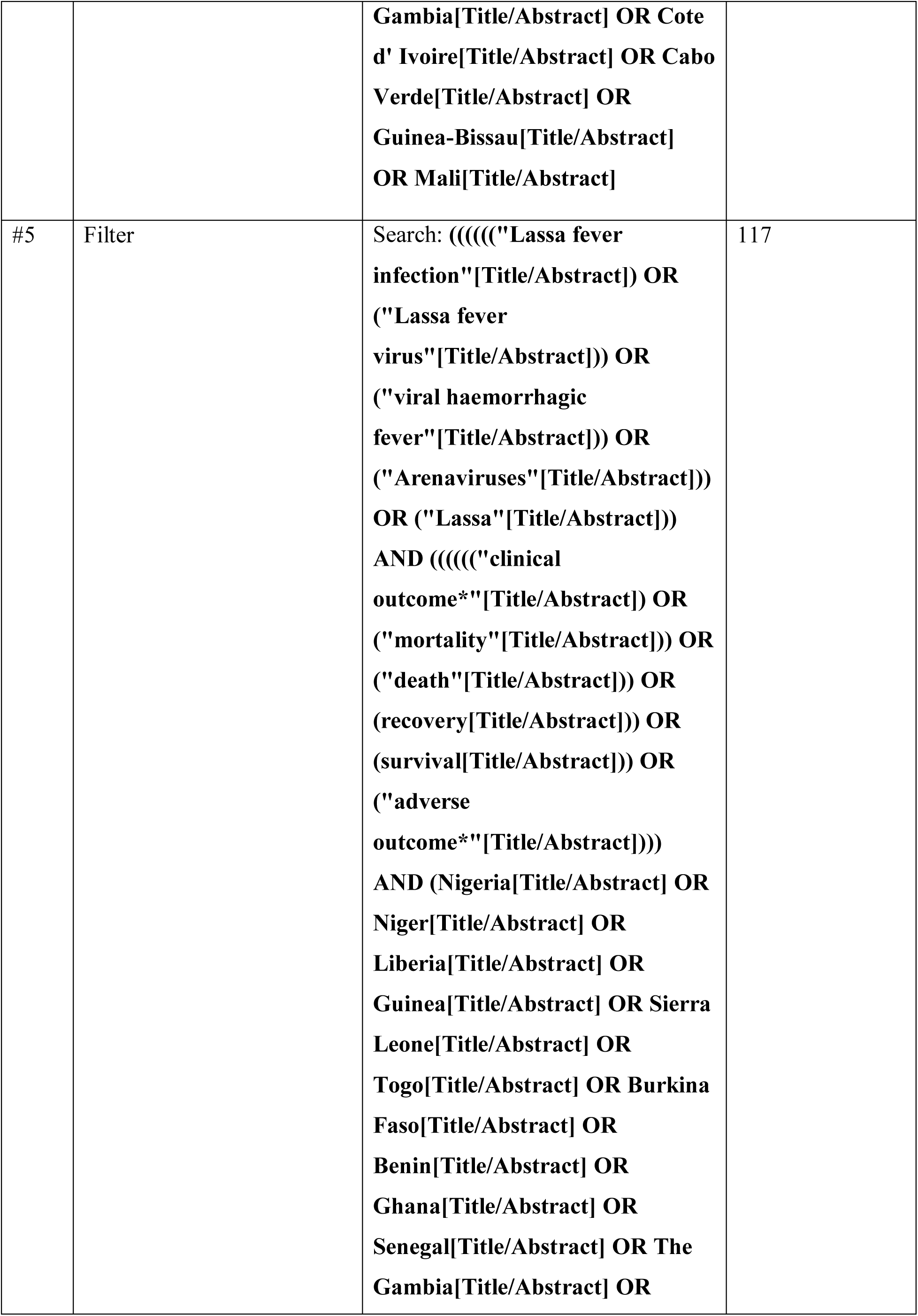
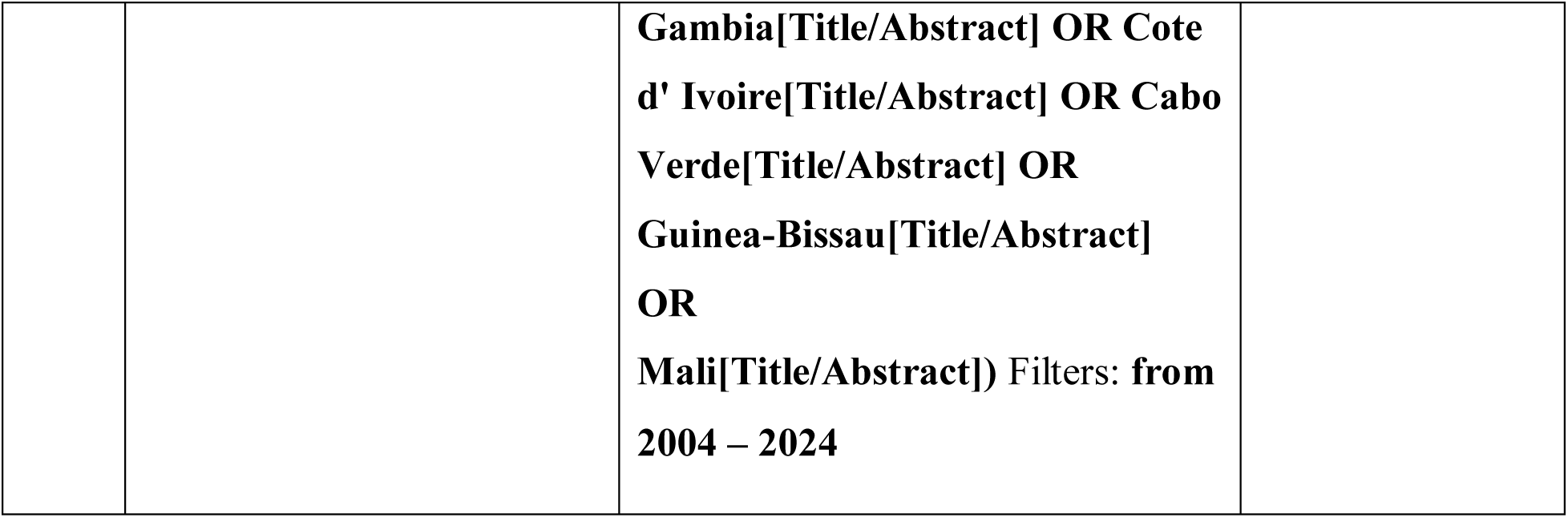
Search Strategy.

